# Integrated Genomic and Transcriptomic Profiling Reveals Distinct Biological Programs and Therapeutic Opportunities in Cancer of Unknown Primary

**DOI:** 10.64898/2026.11.14.26368097

**Authors:** Wenya Wang, Rosan Olsman, Dwin G. B. Grashof, Lidy Scheffers, Michaël Leenders, Debbie G. J. Robbrecht, Petur Snaebjornsson, Peter D. Katsikis, Yes A. J. van de Wouw, Harmen J.G. van de Werken

## Abstract

**Background:** Cancer of unknown primary (CUP) is a metastatic, heterogeneous malignancy accounting for 2-5% of cancer diagnoses globally. Failure to identify the primary site precludes tumor-type-specific therapy and, together with an aggressive clinical course, contributes to poor survival. The biological mechanisms driving these behaviors remain poorly understood. A deeper understanding of CUP biology is therefore essential to improve diagnostic precision and therapeutic strategies. To address this knowledge gap, we comprehensively characterized the multi-omics landscape of CUP and identified biological and potentially therapeutic targets beyond tissue-of-origin (ToO) assignment.

**Methods:** Whole-genome sequencing (WGS) data from 283 CUP patients were analyzed for ToO predictions, mutational signatures, driver alterations, genetic immune escape (GIE), and clinically actionable alterations. Matched RNA-sequencing (RNA-seq) data from 79 cases were used to characterize functional gene expression signatures and define transcriptomic subtypes. Clinical, WGS, and RNA-seq data were integrated via weighted gene co-expression network analysis (WGCNA).

**Results:** High-confidence ToO predictions (likelihood >80%) were obtained for 58% of CUPs, most commonly predicting lung, gastrointestinal, and biliary tract origins. Mutational signatures clustering revealed four subgroups dominated by tobacco smoking, defective DNA mismatch repair (dMMR), UV exposure, and APOBEC activity, while a fifth group, comprising most tumors (n=169; 60%), showed mixed mutational processes. The cohort also frequently exhibited mutations in established cancer driver genes, including *TP53*, *CDKN2A*, *KRAS*, and epigenetic modifiers *ARID1A, SMARCA4, and ARID2*. GIE alterations occurred in 35% of tumors, mostly involving *HLA-I* genes. Transcriptomic profiling defined five subtypes with different degrees of immune infiltration, fibrosis, and tumor proliferation. These subtypes were not significantly associated with clinical or genomic features, indicating that the tumor microenvironment (TME) represents another layer of CUP heterogeneity. WGCNA further linked proliferative programs to cell-cycle, DNA-replication, and neuroendocrine features. WGS data identified potentially targetable alterations in 64% of the CUP patients.

**Conclusions:** CUP comprises biologically heterogeneous tumors with diverse predicted origins, mutational histories, and TME subtypes. Transcriptomic profiling and integrated genomics suggest that immune escape and rapid proliferation contribute to the clinical aggressive behavior. Together, these findings provide a molecular framework for understanding CUP heterogeneity and support therapeutic stratification by both ToO predictions and tumor-specific molecular alterations.

## Introduction

Cancer of unknown primary (CUP) is a clinically and biologically heterogeneous group of metastatic tumors for which the anatomical site of origin remains unidentified despite extensive diagnostic evaluation[1]. While advances in diagnostic technologies have reduced the incidence, CUP still accounts for 2-5% of all newly diagnosed cancers worldwide [2]. In the Netherlands, for example, CUP accounts for 0.4% of all new cancer cases, corresponding to approximately 602 diagnoses in 2025[3]. Because the primary tumor site cannot be identified, CUP poses significant clinical challenges. Treatment for most patients remains largely limited to empiric chemotherapy, as site-specific chemotherapy is not an option, contributing to a poor overall survival of only 6-8 months[4,5]. However, the recent CUPISCO study showed improved outcomes with sequential chemotherapy and molecularly targeted therapy [6]. The clinical complexity, therapeutic uncertainty, and poor prognosis associated with CUP highlight the need for a deeper understanding of its biology.

Advances in molecular profiling technologies, particularly whole-genome sequencing (WGS) and RNA sequencing (RNA-seq), offer opportunities to investigate CUP biology. Although the underlying biological processes driving CUP tumorigenesis remain incompletely understood, multiple hypotheses have been proposed[7]. One hypothesis suggests that CUP represents metastasis from a conventional primary tumor that remains clinically occult because it is indistinguishable from metastatic disease, dormant, or regressed. Based on this concept, several tissue-of-origin (ToO) genomic prediction tools[8–10] have been developed, including the cancer of unknown primary prediction algorithms (CUPPA)[11], which uses positional mutational distributions, 96-trinucleotide single-base-substitution (SBS) context, driver gene profiles, and other genomic features to infer the likely tissue of origin. Likewise, RNA-based and methylation-based[10] classifiers have shown clinical utility by guiding site-specific therapy, and RNA-based classifiers has been associated with improved progression-free survival[12,13]. An alternative hypothesis proposes that CUP may represent a distinct tumor entity, potentially arising from stem-like cells with high metastatic potential and no clear identifiable primary site[14]. This model is in line with transcriptomic studies showing poor correlation between CUP and known cancer types in The Cancer Genome Atlas (TCGA)[7,15,16].

Irrespective of its origin, CUP is typically characterized by poor prognosis[17], driven in part by rapid proliferation and dissemination to distant sites at diagnosis[14]. In many cancer types, the tumor microenvironment (TME) actively contributes to tumor aggressiveness through diverse mechanisms, including angiogenesis, immune evasion, and metabolic reprogramming[18]. Beyond the TME, genomic profiling of CUP can identify clinically actionable alterations that may guide targeted therapies[15,19]. Importantly, tumors with an inflamed, immune-infiltrated TME may be more responsive to immunotherapy, and several CUP case studies have reported favorable outcomes with immunotherapy combined with other treatments [20–22]. Therefore, integrating genomic profiling with TME characterization may provide novel biological insights and help identify therapeutic opportunities for patients with CUP. However, comprehensive multi-omics profiling of CUPs remains limited, as most studies are constrained by small cohort sizes and a lack of integrated genomic and transcriptomic analyses [23,24].

In this study, we characterized the genomic landscape of 283 CUP patients using WGS and identified driver genes and cancer hallmarks. In addition, we obtained matched RNA-seq data from 79 CUP patients to explore the TME and link transcriptomic data to clinical and genomic traits. Overall, our integrated genomic and transcriptomic approach was designed to advance the understanding of CUP biology and support ongoing efforts to improve diagnosis and treatment for CUP patients.

## Materials and Methods

### Patient cohort

This study aimed to characterize the multi-omics landscape of CUP and identify biological and potentially therapeutic features. Therefore, patients diagnosed with CUP were included from five sources: (a) the Algorithmic Cancer Treatment In the Netherlands (ACTIN; METC 19-0446) initiative, (b) the routine diagnostic samples (CORE) from the Netherlands Cancer Institute (NKI), (c) the Center for Personalized Cancer Treatment (CPCT-02; NCT01855477; start from 2011-08-17)[25], (d) the Drug Rediscovery Protocol (DRUP; NCT02925234; start from 2016-08) study[26], and (e) the Whole Genome Sequencing Implementation in standard Diagnostics for Every cancer patient (WIDE; NL68609.031.18) from the NKI[27].

### Clinical data curation

Two clinicians curated the clinicopathological data: one medical oncologist (YvdW) from VieCuri Medical Center and one pathologist (PS) from the NKI. We excluded tumors with a histopathological diagnosis that follows ESMO Clinical Practice Guidelines (2011) (e.g., neuroendocrine tumors, sarcomas, high-grade serous carcinomas, melanomas, germ cell tumors, and mesotheliomas)[28]. Cases labeled as CUP in the HMF database but shown to have a known primary origin on retrospective clinicopathological review were excluded. We also excluded cases with WGS performed to aid the diagnostic work-up. For patients with multiple biopsies, only the first biopsy was included in the study to maintain consistency across samples.

### Whole-genome sequencing data

Detailed characterization of WGS library preparation and processing is provided in the Supplementary Information (SI) document. In short, raw WGS data were processed by the HMF computational pipeline (https://github.com/hartwigmedical/pipeline5).

### Driver genes enriched with non-synonymous mutations

Potential driver genes were identified through the ratio of non-synonymous (dN) and synonymous (dS) mutations using the dNdScv R package[29] (v0.0.1.0) based on the somatic mutations identified by SAGE. Genes with an FDR-adjusted p-value<0.05 were enriched with non-synonymous mutations and considered driver genes.

### Driver genes frequently altered by CNAs

sCNAs identified by GISTIC2 were mapped to genes annotated in GENCODE (release 38) using the GRCh37 reference genome. A minimum overlap of 10 base pairs was required between each significant GISTIC2 peak and a gene annotation. For each significant peak, all genes meeting this overlap requirement were initially assigned to the corresponding peak. When multiple genes overlapped the same peak, genes present in a manually curated cancer driver gene list were prioritized. If no genes overlapped a peak, the nearest GENCODE protein-coding gene was assigned. The manually curated driver gene list integrates data from multiple sources, including COSMIC Cancer Gene Census (v92), IntOGen[30,31], Compendium Cancer Genes (02-2020), Priestley *et al*[25], Dietlein *et al*[32], Martincorena *et al*[29], and Bailey *et al*[33]. The complete list of driver genes is available in the R2CPCT package[34].

### Tumor mutational burden

Tumor mutational burden (TMB) was calculated based on the total sum of SNVs, MNVs, and InDels per megabase over the total number of callable nucleotides in the human reference genome as previously described[35]. A TMB was classified as low (<5), medium (≥5-≤10), or high (>10).

### Mutational signatures

A 96-trinucleotide mutation count matrix was constructed from the identified SNVs relative to the GRCh37 genome. Single base substitution (SBS) mutational signatures from the Catalogue of Somatic Mutations in Cancer (COSMIC; v3.1)[36] were retrieved using the mutationalPattern package[37] (v3.14.0). The optimal nonnegative linear combination of the trinucleotide contexts was fitted to the known COSMIC signatures to derive SBS signatures. The relative contribution of each signature was calculated and signatures contributing less than 5% were grouped into a separate category (Filtered <5%).

### Putative therapeutic targets

Targetable genomic alterations were identified using the ESMO Scale for Clinical Actionability of molecular Targets (ESCAT) database[38]. Only alterations classified as tier I (ready for routine use) or tier II (investigational use) were retained.

### Germline variants

Germline variants were annotated using the VEP with the gnomAD plugin. Variants with a maximum allele frequency <0.005 were retained. Variants were further filtered to those present in the ClinVar database[39] (downloaded 2024-09-16) and classified as pathogenic or likely pathogenic, with expert or guideline-based review status and germline or germline/somatic origin. Only variants associated with cancer-related phenotypes (cancer, tumor, neoplasm) and predicted to have high impact were included.

### Whole-transcriptome sequencing

Detailed characterization of RNA-seq library preparation and pre-processing is provided in the SI document. In short, we used the nf-core RNA-seq pipeline (v3.12.0) [40] to pre-process raw RNA-seq data.

### Tumor microenvironment analysis

To describe the TME in CUPs, we applied a set of knowledge-based functional gene expression signatures (Fges) that represent key immune and stromal components, curated by Bagaev *et al* [41]. Gene set variation analysis (GSVA) was performed using the GSVA R package[42] (v2.2.1) to quantify the expression activity of each signature across CUP patients. The input for GSVA was the VST-transformed gene expression matrix generated by DESeq2. To subtype the CUP TME, we adopted an approach similar to that of Bagaev *et al*. In short, a similarity matrix was constructed by calculating the Pearson’s correlation coefficient between all CUP samples using the GSVA scores. The matrix was then used to generate a weighted graph, where each node represented a sample and the edge weights corresponded to the Pearson’s correlation values between pairs of samples. Louvain clustering was subsequently applied to detect clusters within the graph using igraph R package[43] (v2.2.0). To determine the optimal weight threshold for defining clusters, we checked the number of samples in each cluster. Candidate thresholds that resulted in any cluster containing <5% of samples were excluded. Subsequently, we evaluated several clustering quality metrics to choose the optimal threshold, including Davies-Bouldin index, Calinski-Harabasz index, and Silhouette score.

### Quantification of hallmark pathway and neuroendocrine activity

The activity of 50 hallmark pathways curated in the Molecular Signature Database[44] (MSigDB) was also quantified using GSVA. To further investigate neuroendocrine features in CUP, the Gavish_3Ca_Metaprogram_Epithelial_Neuroendocrine gene set was analyzed using the same approach.

### Immune cell deconvolution with CIBERSORTx

Immune cell composition was analyzed using CIBERSORTx[45] with the LM22 signature matrix in the relative mode. The analysis used transcripts per million (TPM)-normalized count matrix as input. A batch correction was performed using the B mode for removing technical biases between bulk mixture data and the reference signature, as recommended by the CIBERSORTx authors, and the analysis was carried out with 500 permutations to ensure robustness and accuracy.

### Outcome analysis

Treatment response assessment for CUP patients in the CPCT-02 study was performed using the Response Evaluation Criteria in Solid Tumors (RECIST; v1.1)[46] score. For survival analysis, either the date of death (if available) or the date of the most recent RECIST assessment was used as the endpoint. Survival curves were generated using the survival[47] (v3.8.3) and survminer[48] (v0.5.0) R packages.

### Statistical analysis

All statistical analyses were performed using R (v4.5.1) and associated packages. Group comparisons were conducted using the Mann-Whitney U test for continuous variables, and the *χ^2^*-test or Fisher’s exact test for categorical variables. The BH procedure was used to adjust the p-values in the analyses of 1) differential gene expression, 2) pathway over-representation, 3) feature comparison between transcriptomic clusters. A p-value<0.05 or, when applicable, a p-adjusted value<0.05 was considered statistically significant.

## Results

### Clinical and genomic profiling captures the heterogeneity of CUP

A total of 369 tumor samples from patients initially labeled as CUP were included from the Dutch ACTIN, CORE, CPCT-02, DRUP, and WIDE studies (Figure 1A). Following clinicopathological review, 49 cases were excluded because a primary tumor site was known on review and 27 cases because patients were managed according to ESMO Clinical Practice Guidelines (2011) based on histopathological classification[28]. For patients with multiple available, we retained only the first biopsy for the analysis, resulting in the exclusion of 10 additional samples. The final CUP cohort comprised 283 patients, including 139 females and 144 males (Supplementary Table 1), with all tumor samples obtained through needle biopsies.

**Figure 1.**
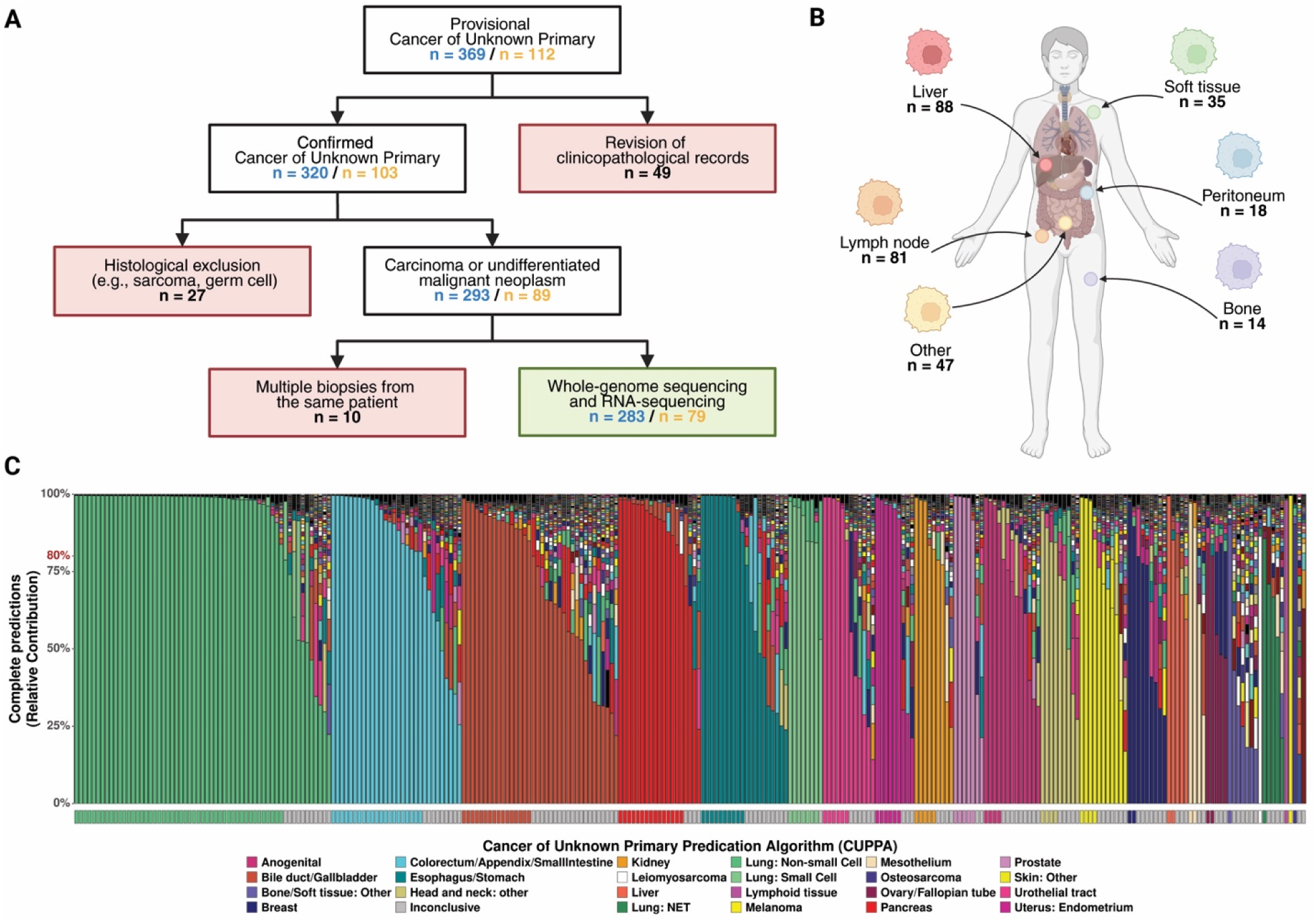
Cancer of unknown primary (CUP) cohort selection, overview, and tissue of origin prediction. (A) Patient inclusion workflow. From 369 CUP cases, clinicopathological review and sample filtering yielded 283 tumor samples from distinct CUP patients. Counts of samples with whole-genome sequencing (WGS; blue) and RNA-sequencing (RNA-seq; yellow) were indicated. (B) Distribution of biopsy sites. Liver, lymph node, soft tissue, peritoneum, and bone were the most frequently sampled metastatic locations. (C) Cancer of unknown primary prediction algorithms (CUPPA) tissue-of-origin predictions. The upper panel shows the relative similarity contribution for all predicted primary types, with the 283 tumor samples on the x-axis and likelihood on the y-axis, and the colors indicate the predicted cancer types. Lower panel highlights high-confidence predictions (>80% similarity threshold value).

Age at biopsy was available for 51 patients from the CPCT-02 and DRUP studies, representing 18% of the total cohort. Among these patients, most were older than 65 years (n=31; 61% of patients with available age data; 11% of the total cohort), followed by patients aged 51-65 years (n=14; 27%; 5% of the total cohort) and 35-50 years (n=6; 12%; 2% of the total cohort) (Supplementary Table1). Biopsies were most frequently obtained from metastatic sites in the liver (n=88; 31%), lymph nodes (n=81; 29%), and soft tissue (n=35; 12%)(Figure 1B). Histopathological analysis revealed diverse histological types, with adenocarcinomas representing the most common subtype (n=136; 48%). In addition, 14 patients (5%) received treatment before biopsy and 48 patients (20%) received treatment after biopsy, predominantly chemotherapy (Supplementary Table 1). Median survival from the date of biopsy was 6 months (Supplementary Figure 1A). Among the 48 patients who received post-biopsy treatment, 38 had first post-treatment RECIST assessments, of whom 8 (21%) had a partial response (PR), 18 (47%) had stable disease (SD), and 12 (32%) had progressive disease (PD). Sequential assessment showed changes in response status over time, including transitions from PR to SD or PD (Supplementary Figure 1B). This poor clinical response was consistent with the aggressive nature of CUP.

Next, we characterized the genomic landscape of the CUP cohort to identify the genomic alterations driving CUP. We processed WGS data from tumor biopsies and matched peripheral blood samples for all 283 patients. The median coverage depth was 38X for germline DNA and 100X for tumor DNA, with a median tumor purity of 0.53 (Supplementary Figures 2A-B). Mutation etiology and cancer type-specific abbreviations can be used to predict tissue of origin (ToO). In addition, the distribution of somatic mutations reflects concordance with DNA methylation and, therefore, chromatin accessibility, which is tissue-specific. These characteristics are implemented in the CUPPA algorithm, which we used to predict ToO from WGS data for all samples. In total, 165 patients (58%) showed a reliable prediction likelihood of >80% for a known primary tumor type. The most frequent predicted tissue origins were non-small cell lung cancer (n=48; 17%), lower gastrointestinal tract (colorectum, appendix, and small intestine; n=21; 7%), and biliary tract (intra- and extrahepatic bile ducts and gallbladder) (n=16; 6%) (Figure 1C).

### Mutational signature analysis revealed major mutagenic processes and genomic instability patterns in CUP

WGS enabled the identification of somatic small mutations ≤50bp and the inference of underlying mutational processes using COSMIC mutational signatures. Hierarchical clustering of mutational signature contributions identified five distinct mutational-signature subgroups within the CUP cohort (Figure 2A; Supplementary Data 1). Four subgroups were dominated by signatures associated with tobacco smoking (n=58; 20%), defective DNA mismatch repair (dMMR; n=14; 5%), UV exposure (n=9; 3%), and APOBEC activity (n=33; 12%). The remaining subgroup, comprising 169 tumors (60%), showed mixed contributions from clock-like, chemotherapy-related, and unknown-etiology signatures (Figure 2B).

**Figure 2.**
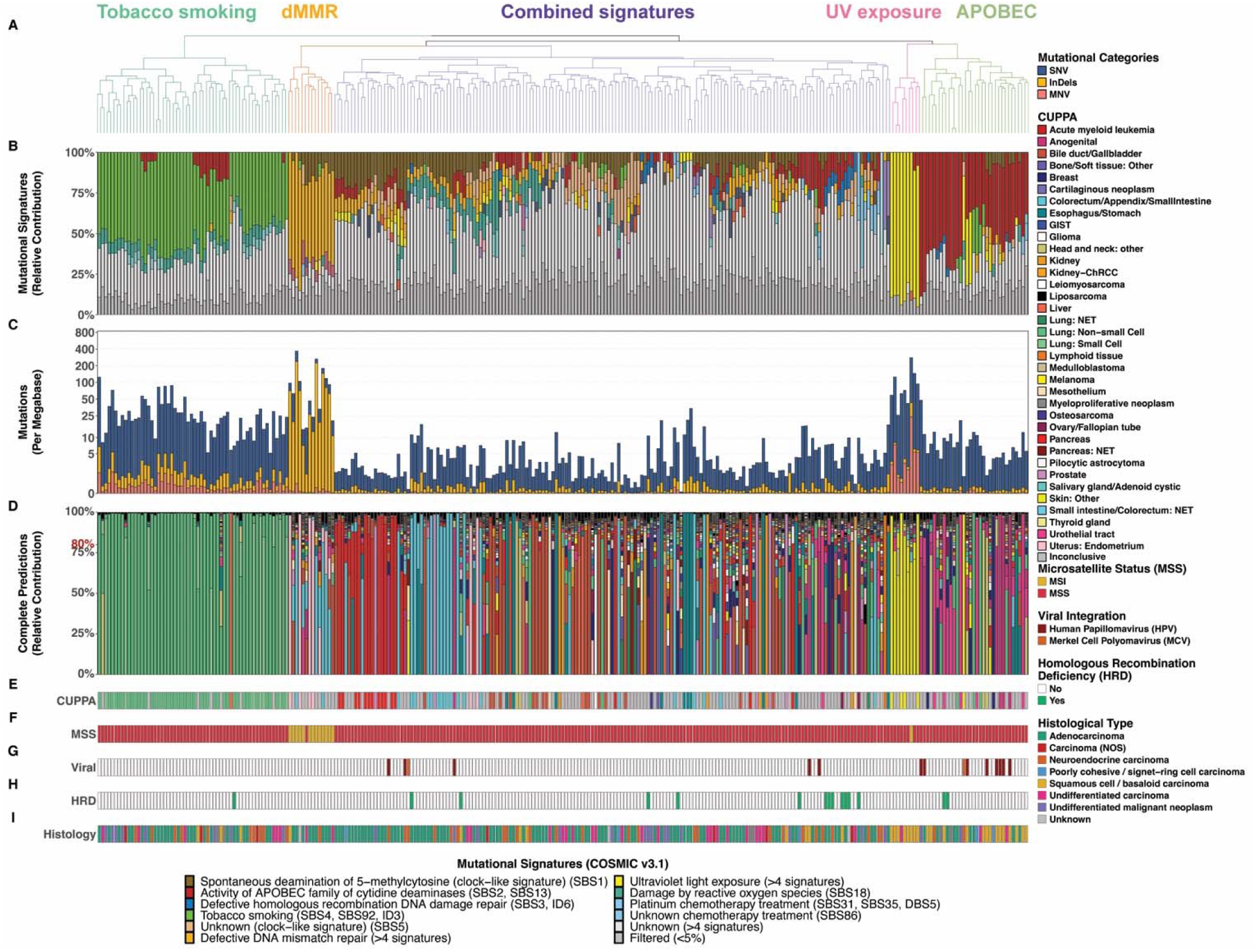
Mutational signatures reveal five distinct genomic subgroups in CUP. (A) Hierarchical clustering of COSMIC mutational signatures identified five genomic subgroups across the CUP cohort, with the 283 samples ordered horizontally. (B) Relative contributions of COSMIC v3.1 signatures per tumor, highlighting dominant etiologies of CUP, which includes tobacco smoking, defective DNA mismatch repair, combined signatures, UV exposure, and APOBEC activity. (C) Tumor mutational burden (TMB) across samples, stratified by mutational categories, including single-nucleotide variants (SNVs), multi-nucleotide variants (MNVs), insertions and deletions (InDels). (D) Relative similarity contribution for all CUPPA-predicted tissues of origin. (E) High-confidence CUPPA predictions (>80% similarity). Samples with no prediction above this threshold were classified as undetermined and colored grey. (F) Microsatellite status, either microsatellite stability (MSS) or microsatellite instability (MSI). MSS in red, MSI in yellow. (G) Detection of viral integrations. Samples with detected viral integration into the genome were colored in red for Human Papillomavirus (HPV) or orange for Merkel Cell Polyomavirus (MCP). (H) Homologous Recombination Deficiency (HRD) status. Samples with detected HRD were colored green. (I) Histological type.

Tumors in the tobacco smoking, dMMR, and UV exposure subgroups were enriched for high TMB, defined as >10 mutations/Mb, consistent with the mutagenic potential of these processes (Figure 2C). All but one tumor in the dMMR subgroup had microsatellite instability (MSI) and a high abundance of insertions and deletions (Figures 2C and 2F). Tumors with a high contribution of UV-induced mutagenesis exhibited a relatively high contribution of multi-nucleotide variants (MNVs), predominantly CC>TT substitutions (Figure 2C). Because CUPPA incorporates mutational-context and other genomic features, ToO predictions were expected to be associated with COSMIC mutational-signature subgroups. Consistent with this, tumors in the tobacco and UV subgroups were frequently assigned lung and skin origins, respectively (Figures 2B, 2D-2E). Evidence of viral integration was detected in 15 of 283 tumors, including Human Papillomavirus and Merkel Cell Polyomavirus (Figure 2G). Another characteristic of genome instability, homologous recombination deficiency (HRD), was detected in 15 tumors (Figure 2H).

Neither viral integration nor HRD was enriched within a specific mutational-signature subgroup or among tumors with conclusive ToO predictions. However, histological type provided context for signature groups. Squamous cell/basaloid carcinomas were significantly enriched within the UV-exposure subgroup (*χ* test, *p*=5.13e-12), providing clinicopathological support for a possible cutaneous origin in a subset of these tumors (Figure 2I).

To further characterize genome instability in the CUP cohort, SVs were analyzed across all tumors (Supplementary Figures 3A-3B; Supplementary Data 2). The median number of SVs was 189, with the first (Q1) and third (Q3) quartiles ranging from 97 to 361, while the relative contribution of different SV types varied substantially between samples (Supplementary Figure 3C). Total SV burden did not differ among the five mutational-signature subgroups, but correlated positively with TMB (Spearman *r*s=0.42; *p*=7.9e-14), ploidy (*r*s=0.22 *p*=2.4e-4), and the number of kataegis foci (*r*s=0.46; *p*=6.7e-16) (Supplementary Figures 3D-3F). Among the mutational-signature subgroups, the APOBEC-enriched subgroup had significantly more kataegis foci than all other subgroups (Kruskal-Wallis test with Dunn’s post-hoc analysis with BH multiple testing correction, adjusted p=6.7e-09), consistent with APOBEC enzymes using single-stranded DNA as substrates. The frequency of chromothripsis, a hallmark of catastrophic chromosomal rearrangement, did not differ significantly among mutational-signature subgroups (Supplementary Figure 3G).

**Figure 3.**
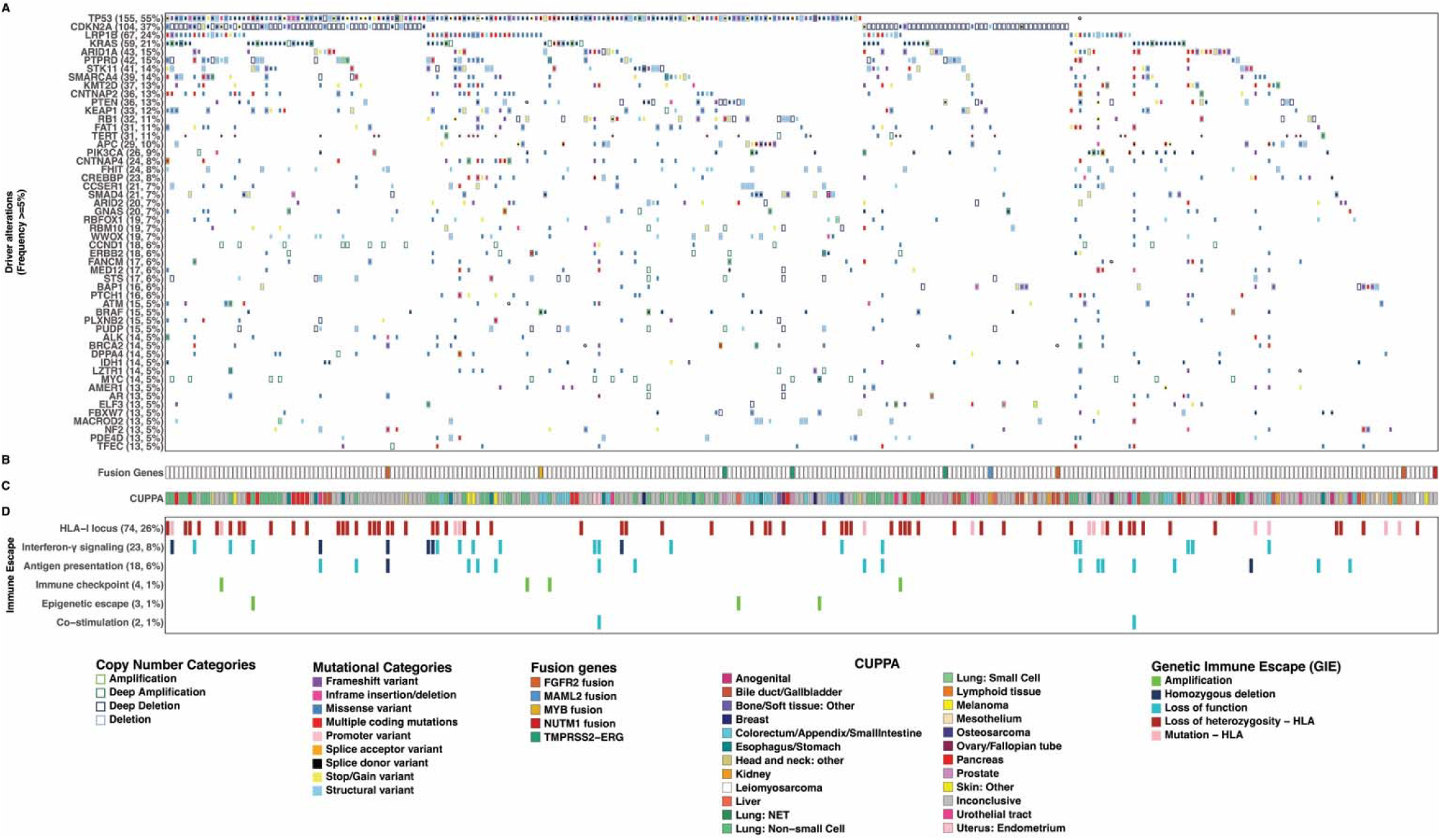
Recurrent driver alterations, fusion events, and immune-escape mechanisms in CUP. (A) Landscape of driver genes that are statistically significant using dN/dS analysis and GISTIC2.0 in ≥5% of tumors that are ordered horizontally. Gene labels include the number of altered cases and their corresponding percentages. Outer boxes represent copy number changes, inner boxes represent small mutations or Structural Variants, boxes marked with a G highlight germline variants, and an asterisk (*) marks hotspot mutations. (B) Presence of known oncogenic gene fusions detected across the cohort. (C) High-confidence CUPPA tissue-of-origin predictions for each sample. (D) Genetic immune escape alterations across six biological pathways, including *HLA-I* locus disruption, antigen presentation, interferon-γ signaling, immune checkpoint genes, epigenetic escape, and co-stimulation pathways.

### Genomic profiling highlights recurrent tumor suppressor loss and immune escape features in CUP

To investigate recurrent genomic alterations with potential functional relevance in CUP, we characterized somatic and pathogenic germline driver alterations, including non-synonymous driver mutations, copy number changes, hotspot mutations, and gene fusions (Figures 3A and 3B). The most frequently altered gene was *TP53* (n=155; 55%), consistent with previous CUP studies [49,50], followed by *CDKN2A* (n=104; 37%), which was predominantly affected by deep deletions (Supplementary Figure 4A). Both genes were canonical tumor suppressors with key roles in cell-cycle regulation. Other recurrently altered genes included *LRP1B* (n=67; 24%), *KRAS* (n=59; 21%), *ARID1A* (n=43; 15%), and *PTPRD* (n=42; 15%), which were involved in oncogenic signaling, chromatin remodeling, and other cancer-associated processes. Significant chromosomal-arm deletions were observed on 9q, 9p, 4q, 4p, 22q, 22p, 18q, 15q, and 19p, whereas 1q, 12q, 12p, and 20q were significantly amplified (Supplementary Figure 4B).

The genes *TP53* (n=89; 31%), *KRAS* (n=53; 19%), and *TERT* promoter variants (n=25; 9%) were most frequently affected by hotspot mutations (Figure 3A; Supplementary Figure 5). We observed pathogenic germline variants in 24 patients (8%). *BRCA2* (n=3; 1%) was the most frequently observed germline variant, followed by *CHEK2*, *MLH1*, *MSH6*, *NTHL1*, and *PMS2*, each identified in two patients, and all were related to DNA-repair pathways (Supplementary Figure 5). We also detected oncogenic fusions annotated in the actionability framework used in this study, including *TMPRSS2-ERG* (n=3; 1%), *FGFR2-AHCYL1* (n=2; 0.7%), and *BRD4-NUTM1* (n=1; 0.4%) (Figure 3B). These detected lineage-informative fusion events, such as *ERG* for prostate cancer and *FGFR2* for cholangiocarcinoma, were concordant with CUPPA predictions (Figure 3C) and were included in the CUPPA training features.

Given the potential role of tumor-immune interactions in CUP biology, we next examined genomic alterations affecting established immune-escape pathways. Overall, 100 patients (35%) harbored mutations in at least one of the six genetic immune escape (GIE)-related pathways (Figure 3D). The most frequently affected category involved the *HLA-I* locus (n=74; 26%), primarily through non-synonymous mutations or loss of heterozygosity, followed by alterations in the interferon-γ signaling (n=23; 8%) and antigen presentation pathways (n=18; 6%). Less frequent events were detected in immune checkpoint (n=4; 1%), epigenetic escape (n=3; 1%), and co-stimulatory (n=2; 1%) pathways (Figure 3D and Supplementary Figure 6A). Tumors harboring GIE mutations exhibited higher TMB compared with those without (*χ*-test; *p*=3.9e-11; Supplementary Figure 6B). In addition, the presence of GIE-related alterations was significantly associated with biopsy site (*χ*-test; *p*=1.41e-09), with lymph node biopsies showing a higher frequency of GIE-related alterations than biopsies from other sites (Supplementary Figure 6C).

### Transcriptomic molecular subtyping of CUP identified distinct immune, stromal and proliferative tumor microenvironment states

To explore immune evasion and the diversity of the tumor microenvironment (TME) in CUP, we analyzed RNA-seq data from 82 samples, of which 79 passed quality control (Supplementary Figures 2C and 2D). In metastatic samples, the bulk transcriptomic profiles may be influenced by including transcripts from healthy tissue of the biopsy site. We, therefore, assessed biopsy site-specific effects on the transcriptome. PCA and *t*-SNE revealed partial clustering according to biopsy site (Supplementary Figure 7) and considered biopsy-site effects when interpreting these expression patterns.

Unsupervised Louvain clustering of the 29 Fges defined five transcriptomic subtypes (Figure 4A): immune-enriched/fibrotic (IE/F; n=9; 11%), immune-enriched/non-fibrotic (IE; n=20; 25%), fibrotic (F; n=24; 30%), immune-depleted/tumor proliferated (D/P; n=19; 24%), and immune-depleted (D; n=7; 9%). These subtypes reflected gradients of immune activation, stromal remodeling, and tumor proliferation (Figure 4B).

**Figure 4.**
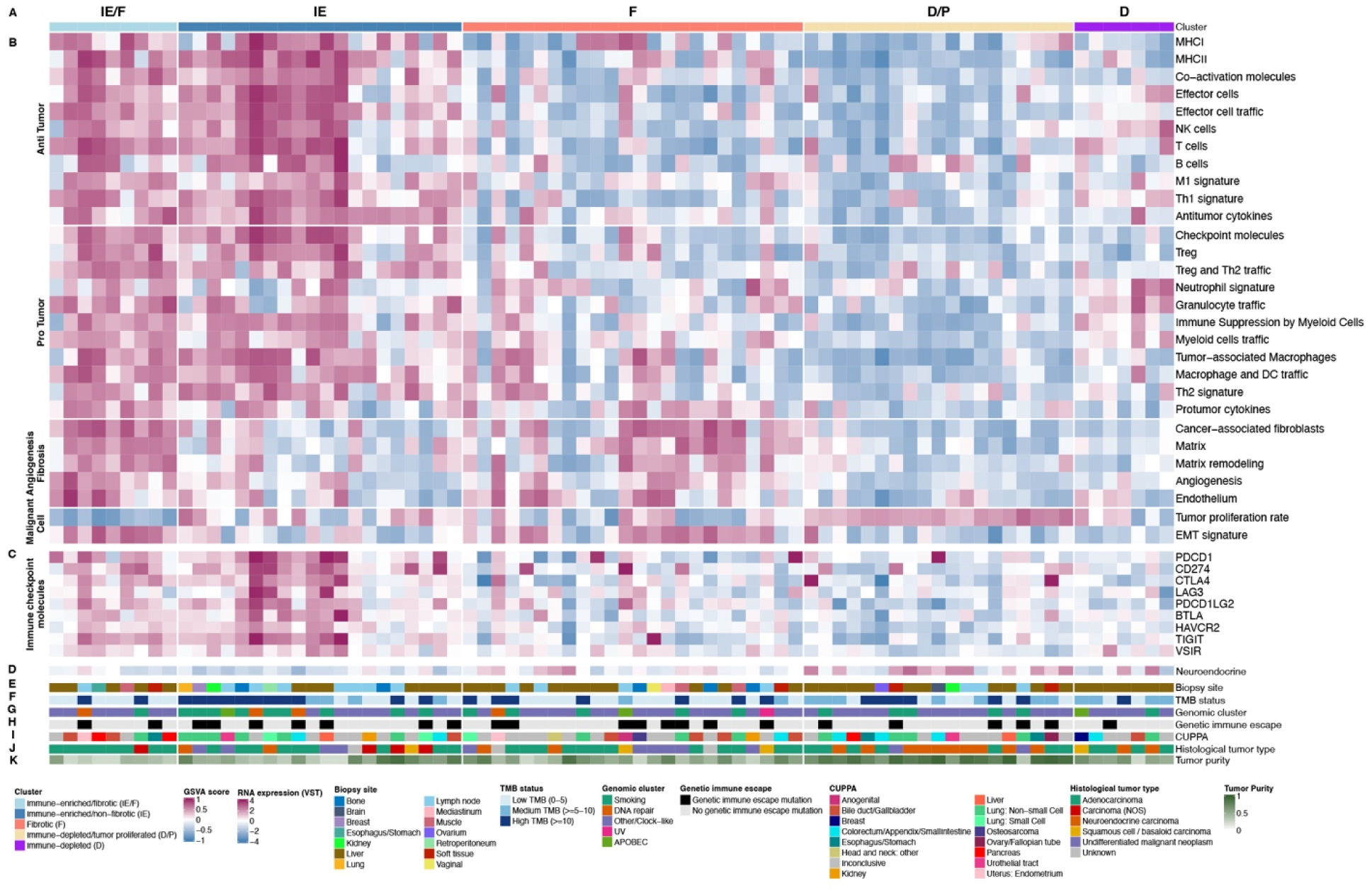
Transcriptomic profiling defines five immune-stromal subtypes in CUP. (A) Louvain clustering of 29 functional gene expression signatures (Fges) identified five transcriptomic subtypes: immune-enriched/fibrotic (IE/F), immune-enriched/non-fibrotic (IE), fibrotic (F), immune-depleted/tumor proliferated (D/P), and immune-depleted (D) for 79 samples ordered on the x-axis. (B) Heatmap of anti-tumor immune cells, pro-tumor immune cells, angiogenesis/fibrosis, and malignant cell property signatures across the five subtypes. (C) Expression levels of nine immune-checkpoint genes. (D) Neuroendocrine-related gene signature expression levels. (E) Biopsy site for each sample. (F) Tumor Mutational Burden (TMB) status. (G) COSMIC multinational signature cluster assignment. (H) Genetic immune escape status. (I) CUPPA tissue-of-origin predictions. (J) Histopathological tumor types. (K) Tumor purity estimates.

The IE and IE/F tumors exhibited strong immune activation and elevated expression of checkpoint genes, including *PDCD1*, *CD274*, *CTLA4*, *LAG3*, and *PDCD1LG2* (Figure 4C). The IE/F subtype additionally showed enrichment of fibrotic and epithelial-organization pathways, including apical junctions and myogenesis signatures, together with upregulation of immune, apoptosis, and p53 programs. This subtype also showed enrichment of oncogenic signaling pathways, including KRAS, Notch, TGF-β, Wnt/β-catenin (Supplementary Figures 8 and 9), suggesting complex immune-stromal interactions. The IE subtype shared interferon responses but was distinguished by increased PI3K/AKT/mTOR signaling (Supplementary Figure 9).

By contrast, the F, D/P, and D subtypes were largely immune-depleted. The F subtype showed fibroblast and ECM remodeling signatures, consistent with stromal activation (Supplementary Figure 8). The D/P subtype demonstrated pronounced expression of a neuroendocrine-associated signature (Figure 4D) together with proliferative programs enriched for cell-cycle, DNA-repair, and mTORC1 pathways (Supplementary Figure 9). At the genomic level, D/P tumors showed a higher burden of SV deletions than other subtypes (adjusted *p*=0.0045) and contained *RB1* mutations in 7 of 19 (37%), although this enrichment was not statistically significant after multiple-testing correction (adjusted *p*=0.25) (Supplementary Figure 10). The D subtypes displayed liver-associated metabolic activity, including bile acid, cholesterol, xenobiotic metabolism, consistent with their biopsy origin of tumors in this group, together with modest neutrophil-related enrichment (Figure 4E and Supplementary Figure 11).

CIBERSORTx analysis supported the subtype assignments, showing higher relative proportions of CD8+ T cells, B cells, NK cells, and M1 macrophages in IE and IE/F tumors than in those of F and D/P (Supplementary Figure 11). No significant subtype differences were observed in driver mutations, genomic subgroups, GIE alterations, CUPPA predictions, or histological tumor types (Figures 4F-4J). Tumor purity differed across subtypes, consistent with variation in cellular composition inferred from the transcriptomic profiles (Figure 4K).

### Clinically actionable alterations are common but context-dependent in CUP

To identify potentially actionable targets and explore therapeutic opportunities beyond empiric chemotherapy, we assessed clinically relevant genomic alterations using the ESCAT framework. Among the 283 patients with CUP, 181 (64%) harbored at least one potentially targetable alteration, with some tumors carrying up to nine such alterations (Figure 5A). A subset of patients had high TMB, a biomarker to support treatment with immune checkpoint inhibitors (ICI)(Figure 5B). Among TMB-high tumors with available transcriptomic subtype information (n=28), 17 (61%) were classified as immune-enriched, including either IE or IE/F subtypes. The remaining TMB-high tumors displayed non-immune-enriched transcriptomic profiles, suggesting that TME context may further refine the interpretation of TMB as a potential immunotherapy biomarker in the context of CUP.

**Figure 5.**
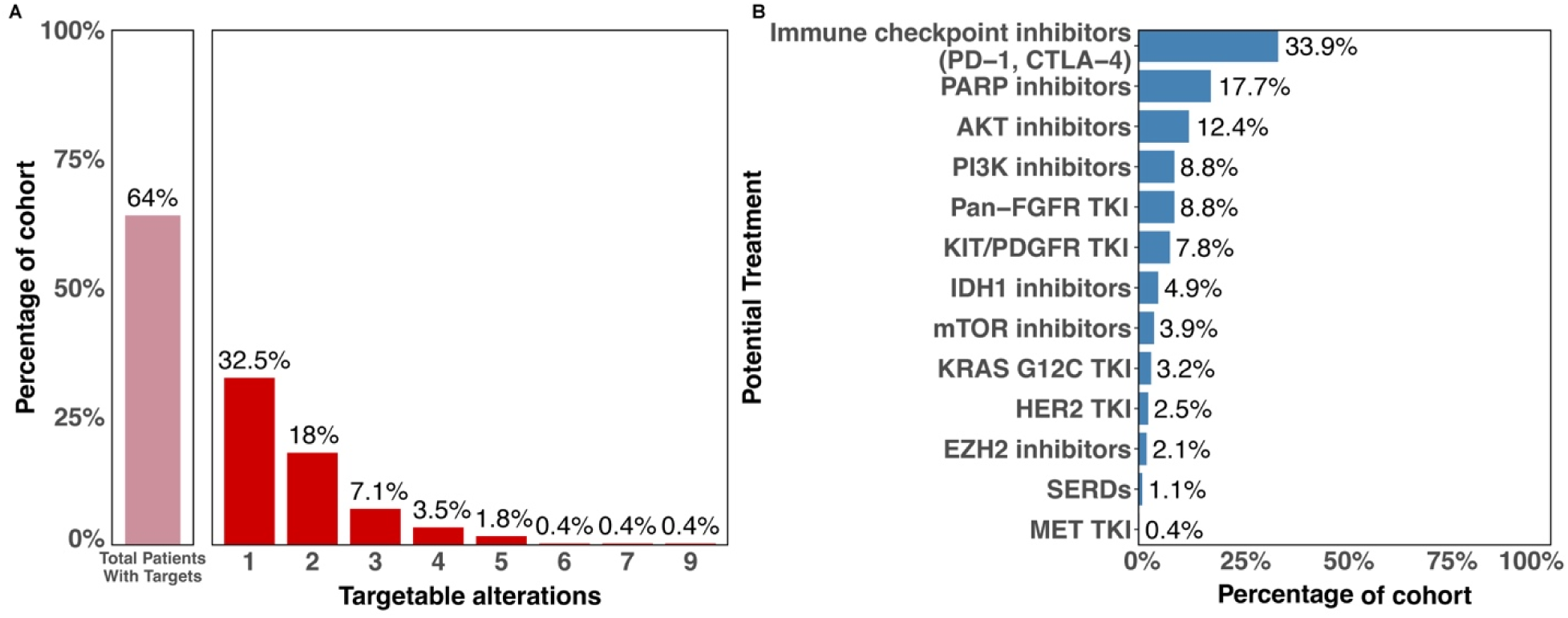
Comprehensive genomic profiling uncovers potentially targetable alterations in the majority of CUP patients. (A) Left bar shows the total number of patients with actionable targets in the CUP cohort; right bar shows the distribution of the number of potentially targetable genomic alterations per patient, shown as percentages of patients with one to nine targetable genomic alterations within the cohort. (B) Potentially targetable genomic alterations for CUP patients based on the ESCAT database.

Actionable alteration-drug matches were identified across several therapeutic classes. FGFR-directed tyrosine kinase inhibitors (TKIs) were linked to tumors harboring FGFR alterations. However, current clinical use of FGFR inhibitors remains largely dependent on tumor type and alteration type rather than tumor-agnostic[51]. Other frequently drug-alteration matches included PARP inhibitors (n=50;18%), mainly associated with ATM mutations or deletions; AKT inhibitors (n=35; 12%), linked to PTEN loss or mutation; and PI3K inhibitors (n=25; 9%), linked to PIK3CA hotspot mutations. KRAS G12C-directed therapy represented an additional potential match in approximately 3% of the cohort (Figure 5B). These assignments indicate molecular indication for treatment rather than demonstrated therapy benefit in this cohort.

### Gene expression network connects key biological and clinical features of CUP

To further integrate transcriptomic programs with clinical and genomic traits of CUP, we performed weighted gene co-expression network analysis (WGCNA) on RNA-seq data. This analysis identified 36 co-expression modules comprising between 37 and 1,814 genes, in addition to a small grey set of three unassigned genes (Supplementary Figure 12). Across the module-trait landscape, notable associations were observed for tumor proliferation, immune and stromal transcriptional signals, CUPPA prediction confidence and biopsy site (Supplementary Figure 13). The tumor proliferation score grouped one set of modules, strongest with the blue module (r_s_=0.91), followed by the pink (r_s_=0.76), turquoise (r_s_=0.45), and red modules (r_s_=0.40; Supplementary Figure 13). These associations should be interpreted cautiously because the proliferation signature partially overlaps with genes comprising these modules. Beyond proliferation, the blue, pink, and red modules were associated with tumor intrinsic characteristics, including higher tumor purity, tumor mutational burden (TMB), *TP53* mutation, and the D/P transcriptomic subtype. The blue, pink, and red modules were also positively associated with neuroendocrine carcinoma histology and negatively associated with adenocarcinoma histology. In addition, the red module was positively associated with the epithelial-neuroendocrine metaprogram (r_s_=0.62).

Functional enrichment using Gene Set Enrichment Analysis (GSEA) further distinguished the biological programs represented by these modules (Supplementary Figure 14). The blue module was primarily characterized by cell-cycle and mitotic processes, including cell-cycle checkpoints, DNA replication and M phase. Consistent with this profile, genes within this module, such as *NDC80[52]* and *CDCA5[53]* (Supplementary Figure 15), have previously been linked with *TP53* mutation or increased *TP53* mutation burden in pan-cancer or tumor-specific analyses. The pink module similarly reflects proliferation activity, with enrichment for DNA replication and replication initiation, chromosome condensation, and origin-of-replication complex assembly. In contrast, the red module was characterized by replication-stress and DNA-repair processes, including ATR activation, homologous recombination and mitotic prometaphase. The turquoise module showed a distinct functional profile, enriched for translation, oxidative phosphorylation, and mitochondrial respiratory electron transport.

A second group of modules, brown, darkorange, black, and darkorange2 captured immune and stromal transcriptional signals (Supplementary Figure 13). Functional enrichment analysis delineated the biological processes represented by these modules (Supplementary Figure 16). The brown module showed a mixed immune-stromal profile, with enrichment of extracellular matrix (ECM) organization and collagen formation alongside cytokine and type I interferon signaling. In contrast, the dark orange module was predominantly associated with immune signaling, including type I and type II interferon pathways and cytokine-cytokine receptor interactions. The black and darkorange2 modules showed dominantly stromal programs, with enrichment of ECM organization, collogen- and proteoglycan-related processes, and integrin signaling. These immune- and stromal-associated modules were also significantly related to our transcriptomic TME subtypes (Supplementary Figure 13).

We next examined the modules correlated with samples for which CUPPA did not yield a confident ToO prediction. Low-confidence CUPPA predictions showed more modest associations with the saddlebrown and darkgrey modules (both r_s_=0.23; Supplementary Figure 13). The darkgrey module was enriched for epigenetic and transcriptional processes, including DNA methylation, histone arginine methylation, and β-catenin/TCF-related transcription, whereas the saddlebrown module showed enrichment for primary-cilium-related processes and RNA polymerase II transcription (Supplementary Figure 17). Both modules were enriched for epigenetic and transcriptional processes, suggesting that tumors with low-confidence genomic ToO predictions may be characterized by altered regulatory states. It remains to be tested whether adding transcriptomic and methylation features would provide complementary information for ToO classification.Finally, given the increased liver-associated metabolic activity observed in transcriptomic subtype D, we investigated whether WGCNA modules were associated with the biopsy site. Indeed, the yellow (r_s_=0.70) and greenyellow (r_s_=0.52) modules correlated with liver biopsy site, and both were characterized by metabolic programs, including cytochrome P450-mediated drug and xenobiotic metabolism, biological oxidation, and retinal metabolism. The yellow module was additionally enriched for complement and coagulation pathways (Supplementary Figure 18). Overall, WGCNA identified distinct co-expression modules associated with proliferation, immune and stromal biology, transcriptional programs associated with low-confidence CUPPA predictions, and liver-associated metabolic activity. Together, these modules highlight coordinated gene-expression patterns across CUP samples that capture complementary aspects of tumor biology, and revealing similar expression patterns across CUP patients.

## Discussion

In this study, we interrogated and integrated whole-genome and transcriptomic sequencing data to characterize the genomic landscape, tumor microenvironment (TME), and potential therapeutic vulnerabilities of CUP. Using WGS data from 283 patients, we identified mutational processes, recurrent driver alterations, structural variations (SVs), and putative genetic immune escape (GIE) events. In a subset of 79 patients with matched RNA-seq data, transcriptomic subtyping revealed distinct immune-enriched/depleted, fibrotic, and proliferative TME states. Together, these diverse findings further emphasized the marked heterogeneity of CUP and support the integration of genomic and transcriptomic data, in addition to tissue-of-origin (ToO) prediction, to better understand the biology of CUP.

Two predominant hypotheses have been proposed to explain CUP biology[54]. The first proposes that CUP represents metastasis arising from a conventional primary tumor that either cannot be distinguished from metastatic lesions or remains clinically occult, potentially due to early dissemination, dormancy or regression. The second proposes that CUP is an independent entity lacking a conventional primary site counterpart. Several observations in our data support the occult primary hypothesis in a substantial proportion of cases. High-confidence CUPPA predictions were obtained for 58% of tumors, and many of these shared genomic characteristics with cancers with known primaries. For example, tobacco- and UV-associated mutational signatures were linked to lung- and skin-like genomic profiles; *TMPRSS2-ERG* and *FGFR2* fusions were detected in tumors with concordant CUPPA predictions; the enrichment of squamous/basaloid histology in the UV-signature group provided clinicopathological support for a cutaneous origin. However, the remaining tumors assigned as “inconclusive” CUPPA predictions should not be interpreted as solid evidence for an independent CUP entity, as current classifiers are limited by the number and constellation of reference categories and features on which they are trained. Additionally, previous work has shown that, no matter whether DNA-, RNA- or methylation-based, ToO approaches are only able to resolve a subset of clinicopathologically unresolved CUPs[10,55]. Moreover, mutations in epigenetic modifiers such as *ARID1A*, *SMARCA4*, and *ARID2* could shift the distribution of somatic mutations in these cancers and reduce the reliability of RNA- or methylation-based ToO detection. Nevertheless, a previous multi-lesion CUP case study identified a high number of fully shared mutations across all 15 metastases, found remarkably similar expression profiles, and observed distinct “galaxy”-pattern evolutionary trajectories in individual CUPs [16]. Taken together, the characterized profiles suggested an occult primary in many cases but did not establish a separate biological CUP entity for the remaining tumors, especially those with low-confidence ToO prediction.

The transcriptomic analyses added another layer of heterogeneity. We identified immune-enriched/depleted, fibrotic, and proliferative states, and these TME states were not significantly associated with other clinical or genomic features. However, weighted gene co-expression network analysis (WGCNA) revealed significant associations between gene modules, histology, and genomics. The blue module, enriched for cell-cycle and mitotic processes, was also associated with *TP53* alterations, higher TMB, and neuroendocrine characteristics. The proliferation-associated modules identified across our CUP cohort were conceptually consistent with experimental CUP models in which stem-like “agnospheres” showed growth-factor-independent proliferation, rapid multi-organ dissemination, and MAP kinase/MYC axis dependence[14]. Of note, transcriptomic analysis was also partially influenced by biopsy site, particularly through liver-associated metabolic programs. These signals may reflect true metastatic-niche adaptation, contamination from normal tissue, or both. Our transcriptomic analysis can not distinguish these possibilities. But other studies have described site-specific effects in metastatic breast[56] and pancreatic cancer[57].

To investigate which genomic mechanisms might enable immune escape in CUPs, we interrogated putative GIE alterations. They were present in 35% of tumors, most often affecting the *HLA-I* locus, followed by interferon-γ signaling and antigen-presentation pathways. Notably, GIE alterations did not define the transcriptomic TME subtypes, suggesting that GIE and the overall immune state are not equivalent. Similarly, although 61% of TMB-high tumors with RNA-seq were immune-enriched, a substantial fraction were not. This indicates that, for CUP, high TMB does not necessarily correspond to an immune-infiltrated TME, warranting caution when applying only TMB as a biomarker for immune checkpoint inhibitor therapy in CUP.

In this study, 64% of patients harbored at least one actionable alteration. This supports the use of comprehensive molecular profiling as a way of generating new treatment options beyond empiric chemotherapy. A recent WGS study that included an overlapping Dutch CUP population similarly reported potentially actionable alterations in 73% of CUP patients and demonstrated diagnostic and treatment utility of WGS[58]. However, the applicability of these targetable alterations is uncertain because many alteration-drug associations depend on the specific combination of alteration plus tumor type. Therefore, further clinical trials are needed to expand the scope of tumor-agnostic alterations and therapies in CUP.

Several limitations should be mentioned. Biopsy site and tumor purity affected bulk transcriptomes, so they cannot fully separate malignant cells from local immune, stromal, and normal tissue components. The cohort also lacked a systematic non-CUP metastatic comparator, limiting research on CUP-specific biology. Finally, the authors inferred actionability from retrospective molecular evidence rather than prospectively testing it.

Future studies should first specifically investigate CUP tumors that lack high-confidence ToO predictions. Matched multi-lesion samples and single-cell or spatial profiling could be used to distinguish tumor-intrinsic signals from environmental adaptation to, or by the metastatic tissue environment. Additionally, methylation profiling may provide complementary information to genomic and transcriptomic features, and cell-free (cfDNA) methylation has already shown feasibility for ToO prediction in CUP[10]. In current inconclusive CUPs, particularly when samples could be collected over time, these approaches would further help determine whether diagnostic uncertainty reflects an unusual ToO or biological changes that occur during tumor progression and metastasis.

### Conclusions

Our integrated genomic and transcriptomic analysis provides a comprehensive, detailed view of the molecular landscape of CUP, revealing the molecular heterogeneity across CUP tumors. While many CUP cases exhibit recognizable mutational features of established cancer types. transcriptomic profiling identified distinct immune, stromal, and proliferative states that are not ACTIN Algorithmic Cancer Treatment In the Netherlands fully captured by genomic subgroups or ToO predictions. These findings demonstrate that ToO assignment alone does not fully reflect the biological complexity of CUP. Integrating genomic, transcriptomic, and epigenomic data may further refine biological stratification and identify therapeutic vulnerabilities, while prospective studies are needed to establish clinical utility of multi-omics profiling.

## Declarations

### Ethics approval and consent to participate

All studies were approved by local institutional medical ethics review committees and conducted in accordance with good clinical practice guidelines and the Declaration of Helsinki’s ethical principles (see M&M). All patients provided written consent for data use and analysis. The Hartwig Medical Foundation (HMF) made clinical, WGS, and RNA-seq data available under data request number DR-196.

### Consent for publication

Not applicable

### Availability of data and materials

The datasets (clinical, WGS, and RNA-sequencing) supporting the conclusions of this article can be freely requested for academic use at HMF (https://www.hartwigmedicalfoundation.nl/en/data/data-access-request/). The HMF pipeline is freely available (https://github.com/hartwigmedical/pipeline5). The code for the analyses and figures in this study is available upon request.

### Author contribution

WW: Writing – original draft; methodology; software; data curation; formal analysis; visualization

RO: Writing – original draft; methodology; software; data curation; formal analysis; visualization

DGBG: Writing – review & editing; methodology; software; formal analysis; visualization

LS: Data curation; writing – review & editing

ML: Data curation; writing – review & editing

DGJR: Data curation; writing – review & editing

PS: Data curation; writing – review & editing

PDK: Writing – review & editing; supervision

YAJW: Supervision; conceptualization; data curation; writing – review & editing; resources

HJGvdW: Supervision; conceptualization; data curation; methodology; formal analysis; data curation; writing – original draft, review & editing; resources

## Acknowledgements

We sincerely thank the Hartwig Medical Foundation for providing the clinical, genomic, and transcriptomic data files used in this study under Data Request Number DR-196. Additionally, we would like to acknowledge the research initiatives and facilities collecting the patient materials from which these data were generated, including the *CPCT consortium*, the diagnostic facility of the Netherlands Cancer Institute, and the DRUP, ACTIN, and WIDE studies. We thank Dr. Floris Groenendijk and Jesse Wolters for their assistance with annotating the clinical records.

## Additional file

Additional file 1

File format: docx (.docx)

Title: Supplementary Information

Description: This file contains supplementary materials and methods, including detailed descriptions of sequencing library preparation, sequencing data processing, and additional methods for analyzing genomic and transcriptomic data. It also contains supplementary table and figures supporting the analyses presented in the main manuscript.

## List of abbreviations

BH: Benjamini-Hochberg
CUPPA: Cancer of unknown prediction algorithms
CUP: Cancer of unknown primary
COSMIC: Catalogue of Somatic Mutations in Cancer
CPCT: Center for Personalized Cancer Treatment
CHORD: Classifier of Homologous Repair Deficiency
dMMR: Defective DNA mismatch repair
DRUP: Drug Rediscovery Protocol
EBV: Epstein-Barr Virus
ESCAT: ESMO Scale for Clinical Actionability of molecular Targets
ECM: Extracellular matrix
FDR: False discovery rate
F: Fibrotic
FC: Fold Change
Fges: Functional gene expression signatures
GSVA: Gene set variation analysis
GIE: Genetic immune escape
GISTIC: Genomic Identification of Significant Targets in Cancer
GRIDSS: Genomic Rearrangement Identification Software Suit
GRIPSS: GRIDSS Post Somatic Software
HMF: Hartwig Medical Foundation
HBV: Hepatitis B virus
HRD: Homologous recombination deficiency
HHV-8: Human Herpesvirus 8
HPV: Human Papillomavirus
ICIs: Immune checkpoint inhibitors
D: Immune-depleted
D/P: Immune-depleted/tumor proliferated
IE/F: Immune-enriched/fibrotic
IE: Immune-enriched/non-fibrotic
InDels: Insertions and deletions
MCV: Merkel Cell Polyomavirus
MSI: Microsatellite instability
MSS: Microsatellite stable
MSigDB: Molecular Signature Database
MNVs: Multi-nucleotide variants
NKI: Netherlands Cancer Institute
PCA: Principal component analysis
PURPLE: Purity Ploidy Estimator
RECIST: Response Evaluation Criteria in Solid Tumors
RNA-seq: RNA-sequencing
SBS: Single base substitution
SNVs: Single-nucleotide variants
SAGE: Somatic Alterations in Genome
sCNAs: Somatic copy number alterations
SV: Structural variants
t-SNE: t-distributed stochastic neighbor embedding
TCGA: The Cancer Genome Atlas
ToO: Tissue-of-origin
TIN: Transcript integrity number
TPM: Transcripts per million
TME: Tumor microenvironment
TMB: Tumor mutational burden
TKIs: Tyrosine kinase inhibitors
VST: Variance-stabilizing transformation
VEP: Variant Effect Predictor
WGCNA: Weighted gene co-expression network analysis
WIDE: Whole genome sequencing Implementation in standard Diagnostics for Every cancer patient
WGD: Whole-genome duplication
WGS: Whole-genome sequencing

