## Supplementary Information for "Integrated Genomic and Transcriptomic Profiling Reveals Distinct Biological Programs and Therapeutic Opportunities in Cancer of Unknown Primary"

Materials & Methods, Tables & Figures

### Supplementary Materials and Methods

#### Whole-genome sequencing

##### Library preparation and data pre-processing

DNA from fresh-frozen tumor tissue and peripheral blood were extracted using the QIAsymphony DSP DNA Mini kit and DSP DNA Midi kit according to the manufacturer’s instructions (Qiagen), respectively, as described previously[1]. Libraries were prepared for a total of 50-200 ng of DNA using the TruSeq Nano LT (Illumina) library preparation prior to sonication into fragment lengths of 450 nucleotides. The HiSeqX platform (Illumina) was used to sequence paired-end reads of 2x150 bases. Alignment and pre-processing of the data were performed by HMF using their WGS analysis pipeline (https://github.com/hartwigmedical/pipeline5). In short, read mapping was performed against the reference genome GRCh37 using BWA-mem[2] (v0.7.17). The mean read coverage for WGS was calculated using Picard[3] (CollectWgsMetrics; v3.8.0).

##### Somatic variant calling, annotation, and filtering

Somatic single-nucleotide variants (SNVs), multi-nucleotide variants (MNVs), and insertions and deletions (InDels) were identified using the HMF in-house Somatic Alterations in Genome (SAGE; v3.2.3) tool in the WGS analysis pipeline, by comparing tumor tissue with the matched peripheral blood reference. Somatic variants were annotated with the Ensembl Variant Effect Predictor[4] (VEP; v104). Variants that failed the default quality control of the Purity Ploidy Estimator (PURPLE; v3.8.4) or had an allele frequency greater than 0.001 in the gnomAD exome database or 0.005 in the gnomAD genome database were excluded.

Structural variants (SVs) were called, filtered, and annotated by HMF using the Genomic Rearrangement Identification Software Suite (GRIDSS; v2.13.2), GRIDSS Post Somatic Software (GRIPSS; v2.3.5), Linx (v1.23.2) and PURPLE. SVs that passed the default PURPLE quality control analyses were retained.

##### Tumor purity, ploidy, and somatic copy number alterations

Tumor purity estimation, ploidy and copy number calling were performed using PURPLE (v.3.8.4) as previously described[1]. In short, PURPLE integrates the B-allele frequency (AMBER), read depth (COBALT), and structural variants to estimate tumor purity, ploidy and copy numbers. In our study, cohort-wise significantly altered somatic copy number alterations (sCNAs) were determined using The Genomic Identification of Significant Targets in Cancer (GISTIC)2.0[5] based on the GRCh37 reference genome. All regions with a false discovery rate (FDR)-adjusted p-value <0.1 were considered statistically significantly altered.

##### Hotspot mutations

Hotspot mutations were assigned by SAGE during somatic variant calling when variants matched a predefined list of known hotspot mutations curated by HMF.

##### Chromothripsis

ShatterSeek[6] (v1.1) was used to identify chromothripsis events. SVs, excluding single-event SVs and those located on the Y chromosome, were used to construct the ShatterSeek SV object. Copy number data, excluding regions on the Y chromosome, were similarly used to build the ShatterSeek copy number object. Both objects were used as inputs for the detection of chromothripsis. A significant chromothripsis event was defined if: (a) ≥25 SVs were present within an event; (b) the maximum number of oscillating copy number segments was≥7 (two states) or the maximum number of oscillating copy number segments was ≥14 (three states); (c) the region of chromothripsis is ≥20Mb; (d) p-value ≤0.05 for the chromosomal breakpoint enrichment test; (e) p-value≤0.05 for the random distribution of breakpoint test.

##### Homologous recombination deficiency

Homologous recombination deficiency (HRD) was detected using the Classifier of Homologous Repair Deficiency[7] (CHORD; v2.3). Only somatic SNVs and InDels, along with all structural variants excluding inversions and single-event SVs, were used as input for CHORD. Samples with an HRD probability≥0.5 were classified as HR-deficient.

##### Microsatellite instability

Microsatellite instability was identified by MSIseq[8] in the HMF pipeline. Tumor samples with an MSI score≤4 were classified as microsatellite stable (MSS), otherwise were classified as having microsatellite instability (MSI)

##### Kataegis

The Katdetectr[9] R package was used to identify regions of localized hypermutation (kataegis). A genomic segment was classified as a kataegis locus if the mean intermutation distance was ≤1000 base pairs and it contained ≥6 somatic variants.

##### Virus detection

Viral integration was detected using VIRUSBreakend (v2.11.1) [10] in the HMF pipeline and subsequently annotated with the HMF in-house Virus Interpreter, focusing on the following species taxa: Human Papillomavirus (HPV), Epstein-Barr Virus (EBV), Merkel Cell Polyomavirus (MCV), Hepatitis B Virus (HBV), or Human Herpesvirus 8 (HHV-8).

##### CUPPA prediction

The Cancer of Unknown Primary Prediction Algorithm[11] (CUPPA; v1.8.1) within the HMF pipeline was used to infer the potential primary site of each CUP based on genome-wide mutation location, 96-trinucleotide mutational context, fusion genes and other driver events, , structural variants, and viral insertions, TMB and whole-genome duplication (WGD). Samples with a CUPPA likelihood score >80% were considered high-likelihood predictions and were assigned to the corresponding predicted cancer type.

##### Genetic immune escape

Genetic immune escape (GIE) mutations were identified by integrating HLA-typing (LILAC; v1.4), small somatic mutations (SAGE) and sCNAs (PURPLE), following the approach of Martínez-Jiménez *et al*.[12]. Six GIE-related pathways were assessed: HLA-I, antigen presentation, interferon-γ, PD-L1 receptor, CD58 receptor, and epigenetic immune escape. Pathways were classified as altered based on characteristic inactivation events (e.g., loss of heterozygosity, homozygous deletions, and clonal loss-of-function) or activating gene amplification (e.g., *PD-L1*, *SETDB1*).

#### Whole transcriptome sequencing

##### Library preparation and data pre-processing

RNA-seq was performed on tumor samples from 82 CUP patients. RNA was extracted from the same fresh-frozen tumor tissues used for WGS using QIAsymphony RNA kit, following the manufacturer’s instructions. Libraries were prepared from 100 ng of RNA using the KAPA RNA HyperPrep with RiboErase (HMR) for ribosomal RNA deletion. Sequencing was performed on two Illumina platforms: the NextSeq 550 (paired-end reads of 2x75 bases) and the NovaSeq 6000 (paired-end reads of 2x150 bases). The nf-core RNA-seq pipeline (v3.12.0) [13] was employed for quality control (FastQC v0.11.9), trimming (TrimGalore! v0.6.7), alignment (STAR v2.7.9a) and quantification (Salmon v1.10.1) against the GRCh38 reference genome and GENCODE annotation (v44). Additional quality assessments, including alignment metrics and transcript integrity number (TIN), were conducted using RSeQC (v3.0.1).

##### RNA-seq data filtering and dimensionality reduction

RNA samples with a TIN value >60 were included for downstream analysis. Principal component analysis (PCA) and *t*-distributed stochastic neighbor embedding (*t*-SNE) were performed to identify variation and detect potential outliers. Genes with fewer than 10 counts across all samples were excluded. The raw gene counts were first transformed using the variance-stabilizing transformation (VST) from DESeq2[14] (v1.45.3). The top 30% of the most variable protein-coding genes after VST were used as input for the *t*-SNE and PCA. The *t*-SNE analysis was conducted with the Rtsne package[15] (v0.17), using a perplexity value of 20. PCA analysis was conducted and visualized using the plotPCA function from DESeq2.

##### Identification of biopsy-specific highly expressed genes

To mitigate potential confounding effects, differential expression analysis was conducted to identify biopsy/tissue-specific highly expressed genes that could potentially introduce bias in the analysis by expressing genes in normal cells from the biopsy site tissue. Comparisons were made between tumors from the liver (n=42), lymph node (n=15), and bone (n=5) against all other tumor samples using DESeq2. Genes with a log_2_ Fold Change (log_2_FC)>1, baseMean>100, and Benjamini-Hochberg (BH) corrected p-adjusted value <0.05 were labeled as biopsy-specific highly expressed genes. Differential expression analysis identified 797 biopsy-site-enriched genes, of which 22 overlapped with the 29 Fges used for subtyping, including *HLA-DQB1* and *TNFRSF9*. For the TME analysis, these overlapping genes were retained because of their established relevance to the TME.

#### Gene co-expression network analysis

Weighted gene co-expression network analysis[16] (WGCNA; v1.73) was performed using VST-transformed RNA-seq data from protein-coding genes with a mean expression>10 counts, which served as the input for the analysis. The absolute Spearman’s correlation coefficient was used to construct an unsigned gene similarity matrix, which was then transformed to a weighted adjacency matrix based on the soft threshold value (β). The optimal β was determined when the correlation coefficient for scale-free topology fitting was >0.85. In our study, a β of five was chosen as the best value for soft thresholding. Modules were defined with a minimum size of 30 co-expressed genes. Module eigengenes were correlated using Spearman’s correlation coefficient to multiple clinical, genomic and transcriptomic traits and p-values were estimated using the asymptotic Student’s *t*-approximation. Genes within modules were further examined with gene set enrichment and overrepresentation analyses against MSigDB. Modules with an absolute correlation coefficient greater than 0.4 and a p-value less than 0.05 for a given trait were selected for further interpretation.

### Supplementary Table and Figures

#### Supplementary Table 1. Clinical and histopathological characteristics of the CUP cohort

Summary of demographic features, tumor histopathology, and treatment status for all 283 patients included in the CUP cohort. Age information was available for a subset of patients (n=51). ^*^Two patients were pre-treated with multiple-line therapy. Not Available (NA).

| **Characteristics of total CUP cohort** | | |
| --- | --- | --- |
| **Sex** | Female | 139 (49.1%) |
|  | Male | 144 (50.9%) |
| **Age** | <35 | 0 |
|  | 35-50 | 6 (2.1%) |
|  | 51-65 | 14 (4.9%) |
|  | >65 | 31 (11.0%) |
|  | NA | 232 (82.0%) |
| **Histopathological tumor type** | Adenocarcinoma | 136 (48.1%) |
|  | Neuroendocrine carcinoma | 38 (13.4%) |
|  | Squamous cell/basaloid carcinoma | 33 (11.7%) |
|  | Undifferentiated carcinoma | 31 (11.0%) |
|  | Undifferentiated malignant neoplasm | 24 (8.5%) |
|  | Carcinoma (NOS) | 7 (2.5%) |
|  | Poorly cohesive signet cell carcinoma | 5 (1.8%) |
|  | NA | 9 (3.2%) |
| **Pre-treatment** | Chemotherapy | 10 (3.5%) |
|  | Hormonal therapy | 2 (0.7%) |
|  | ^*^Chemotherapy (1^st^), Nuclear therapy (2^nd^) | 1 (0.4%) |
|  | ^*^Chemotherapy and Targeted therapy (1^st^), Chemotherapy (2^nd^) | 1 (0.4%) |
|  | Immunotherapy | 0 (0%) |
|  | Treatment naive | 49 (17.3%) |
|  | NA | 220 (77.7%) |
| **Post-treatment** | Chemotherapy | 42 (14.8%) |
|  | Hormonal therapy | 2 (0.7%) |
|  | Immunotherapy | 2 (0.7%) |
|  | Targeted therapy | 2 (0.7%) |
|  | Treatment naive | 12 (4.2%) |
|  | NA | 223 (78.8%) |

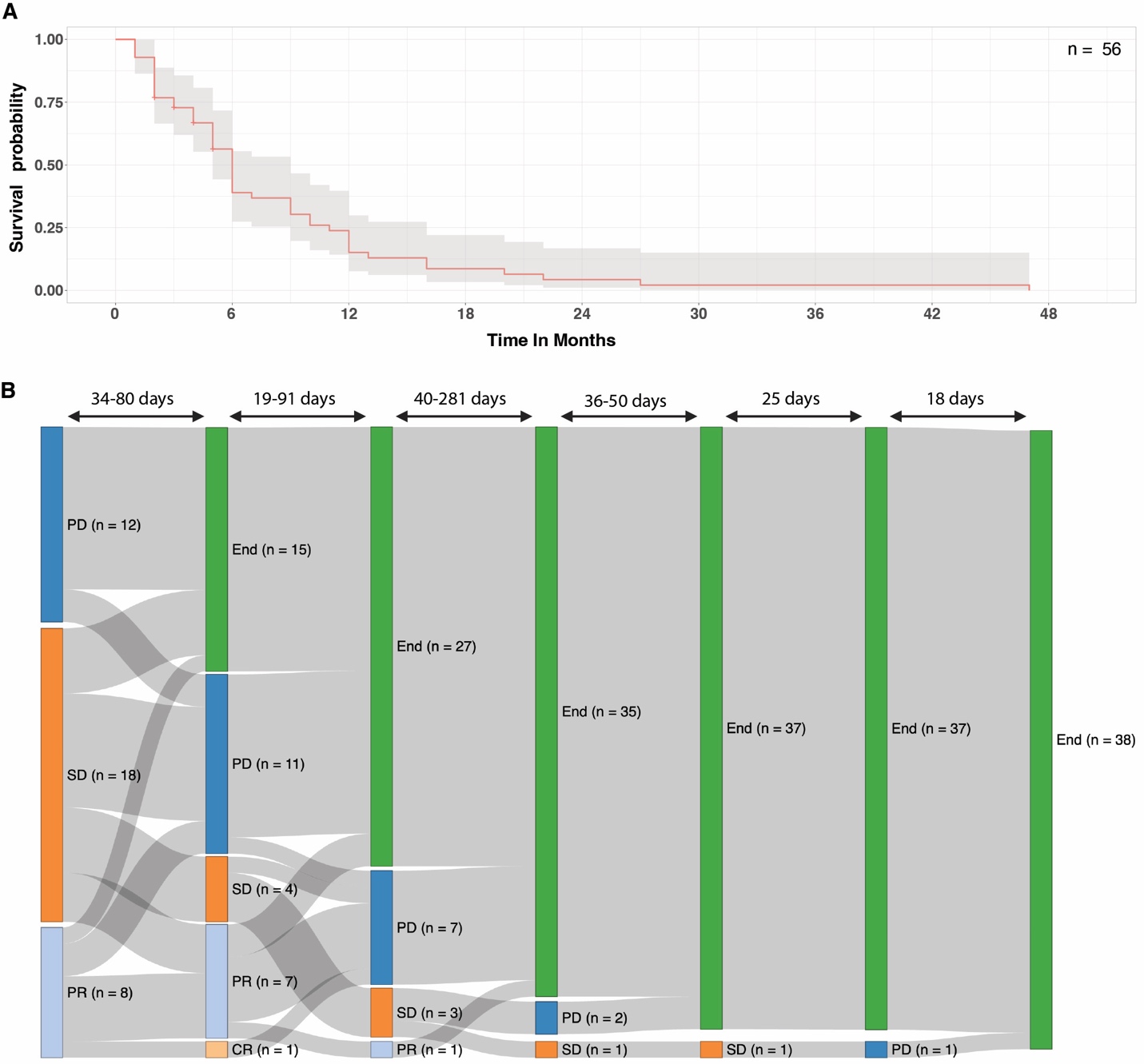

#### Supplementary Figure 1. Poor overall survival and limited treatment responses in the CUP cohort

(A) Kaplan-Meier curve showing overall survival for CUP patients with available follow-up (n=56 with a known date of death or last treatment response date).

(B) Treatment trajectories and response evaluation criteria in solid tumors (RECIST) responses among patients who received post-biopsy therapy (n=38). Each patient had between one to six recorded response evaluations. The number at the top of the diagram indicates the number of days between consecutive RECIST assessments. The label End marks the absence of a subsequent RECIST score (i.e., discontinuation of assessment). CR: Complete Response; PR: Partial Response; SD: Stable Disease; PD: Progressive Disease.

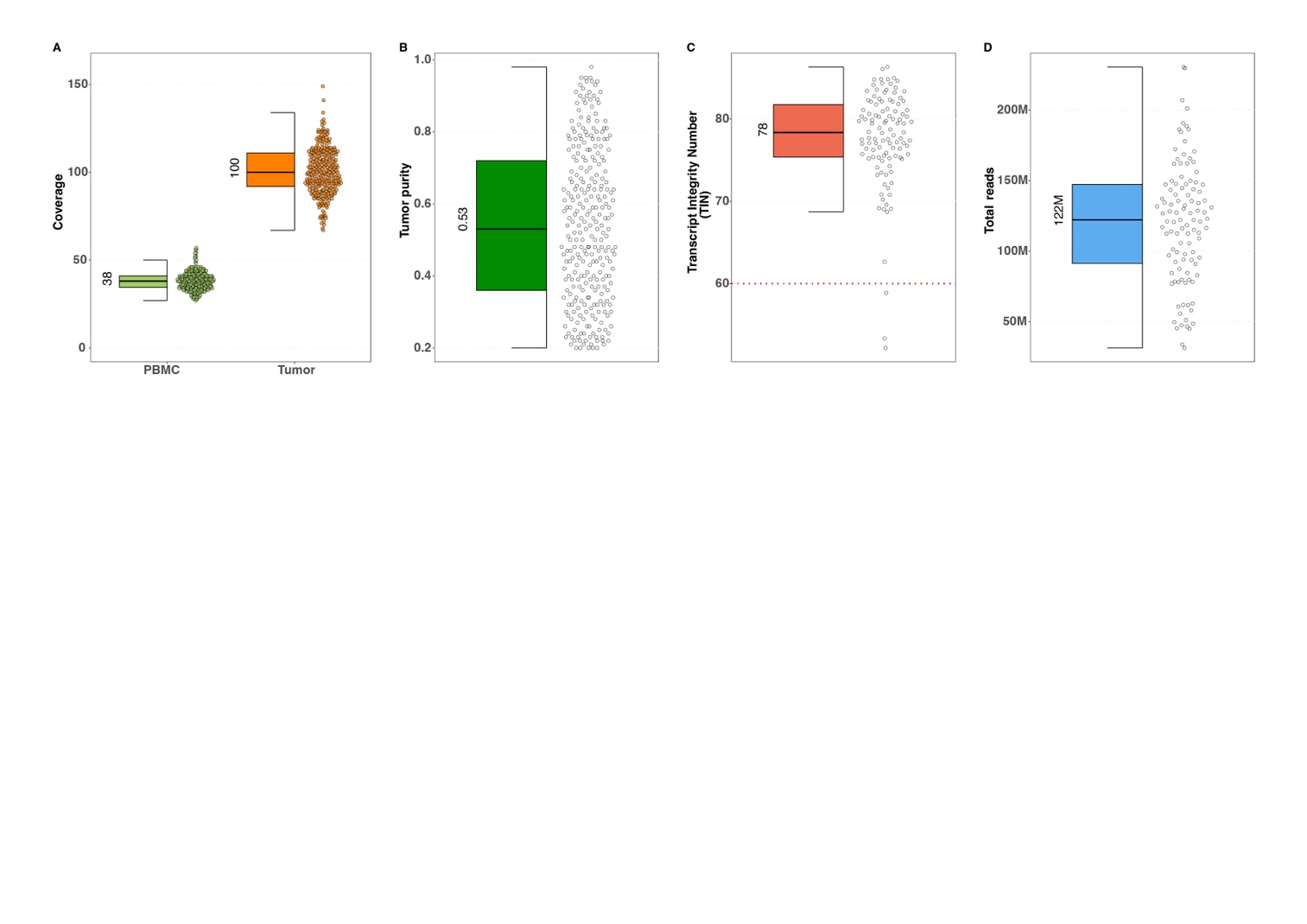

#### Supplementary Figure 2. Sequencing quality metrics for whole-genome sequencing (WGS) and RNA-sequencing (RNA-seq) data in the CUP cohort

(A) Genomic coverage depth of peripheral blood mononuclear cell (PBMC) and tumor WGS samples. Median coverage was 38X for PBMC and 100X for tumor DNA.

(B) Distribution of tumor purity estimates across all WGS samples.

(C) Transcript integrity number (TIN) for RNA-seq samples. 79 of 82 samples passed quality control (QC) with a TIN higher than 60.

(D) Total RNA-seq read counts per sample (paired-end, one pair counted as two reads)

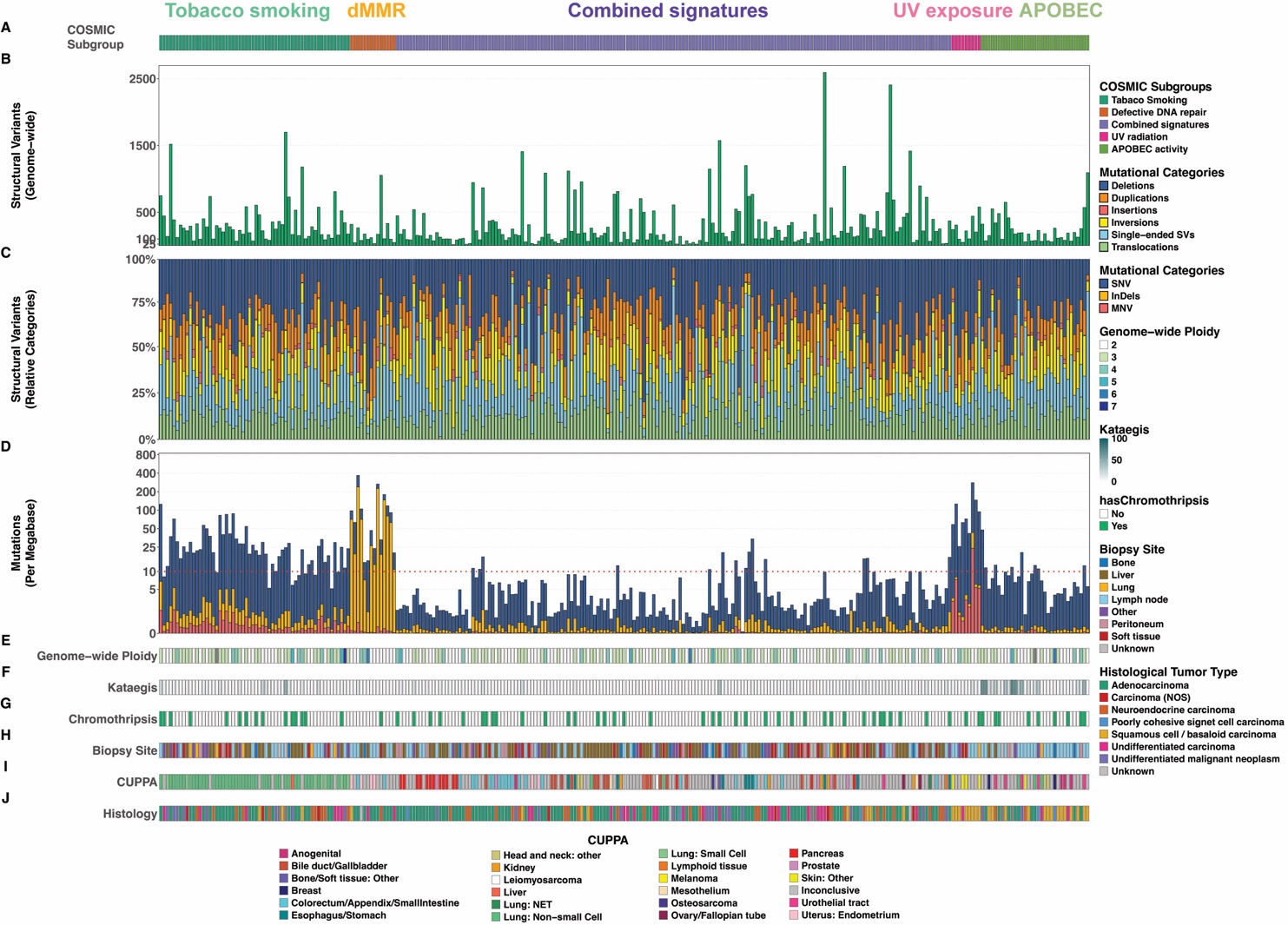

#### Supplementary Figure 3. Structural variant burden and genome instability features across COSMIC genomic clusters

(A) COSMIC mutational signature-based genomic subgroup assignments, following the same sample horizontal order using hierarchical clustering on the COSMIC mutational signatures.

(B) Total number of structural variants (SV) per tumor.

(C) Relative contributions of SV categories, including deletions, duplications, inversions, insertions, single-ended SVs, and translocations.

(D) Tumor mutational Burden (TMB) across samples, stratified by mutational categories: Single Nucleotide Variants (SNVs), Multi Nucleotide Variants (MNVs), and InDels.

(E) Genome-wide ploidy estimates per sample.

(F) Number of kataegis foci per tumor.

(G) Presence or absence of chromothripsis events.

(H) Tumor biopsy site for each sample.

(I) CUPPA tissue-of-origin predictions.

(J) Histopathological tumor classification.

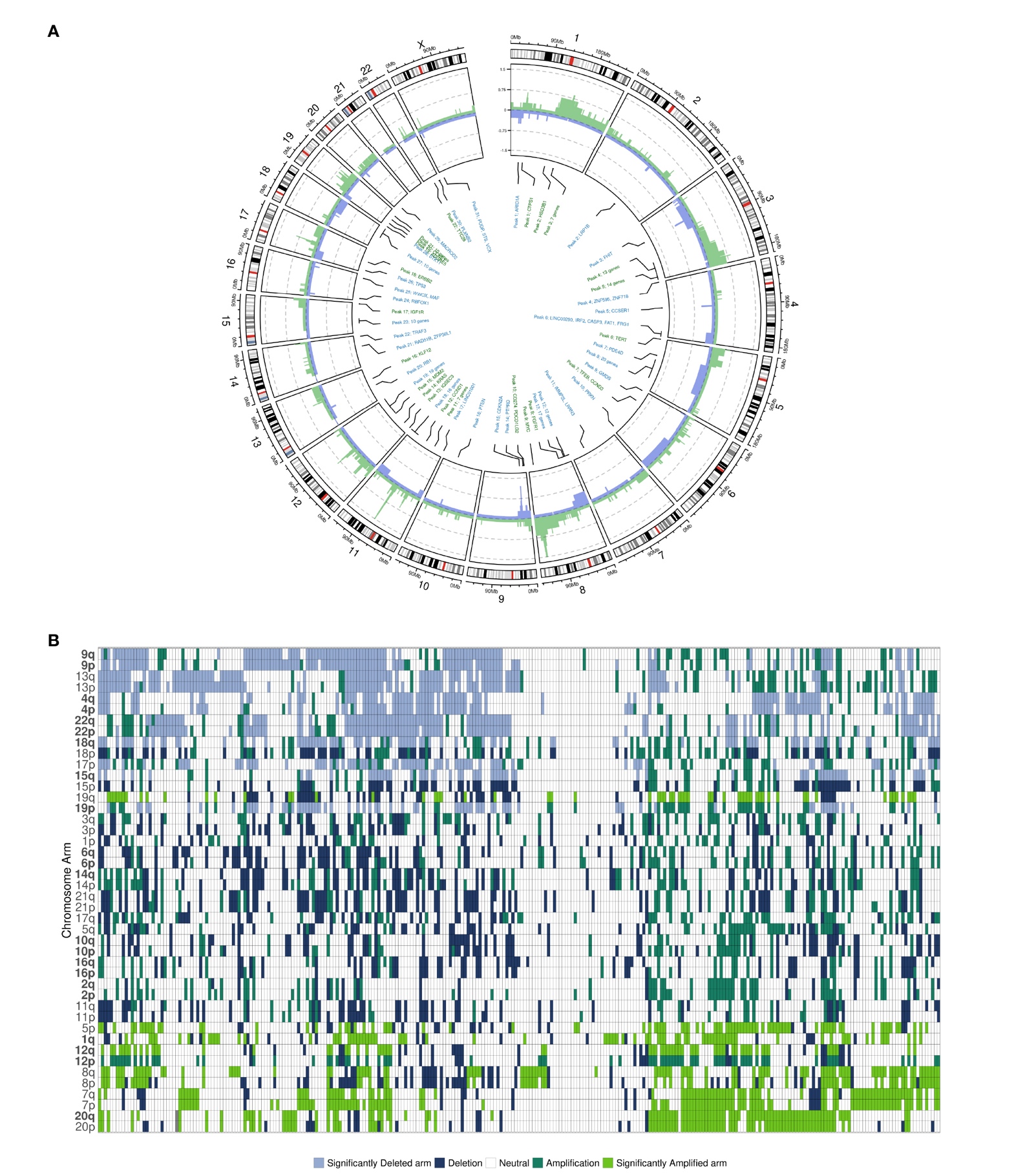

#### Supplementary Figure 4. Genome-wide copy-number landscape of CUP tumors

(A) Circular plot summarizing genome-wide copy number alterations across the CUP cohort. Outer track indicates genomic ideogram. Green indicates amplifications and blue indicates deletions; recurrent alterations and their driver genes are highlighted by name and colored similarly in the inner track.

(B) Arm-level copy number alterations showing deletions and amplifications across all samples (columns). Highlighted chromosome arms (rows) in bold font show statistically significant arm-level changes. Deleted and amplified chromosomal arms are marked in blue and green, respectively. The samples and chromosome arms were clustered using Ward.D2 hierarchical clustering based on Euclidean distance.

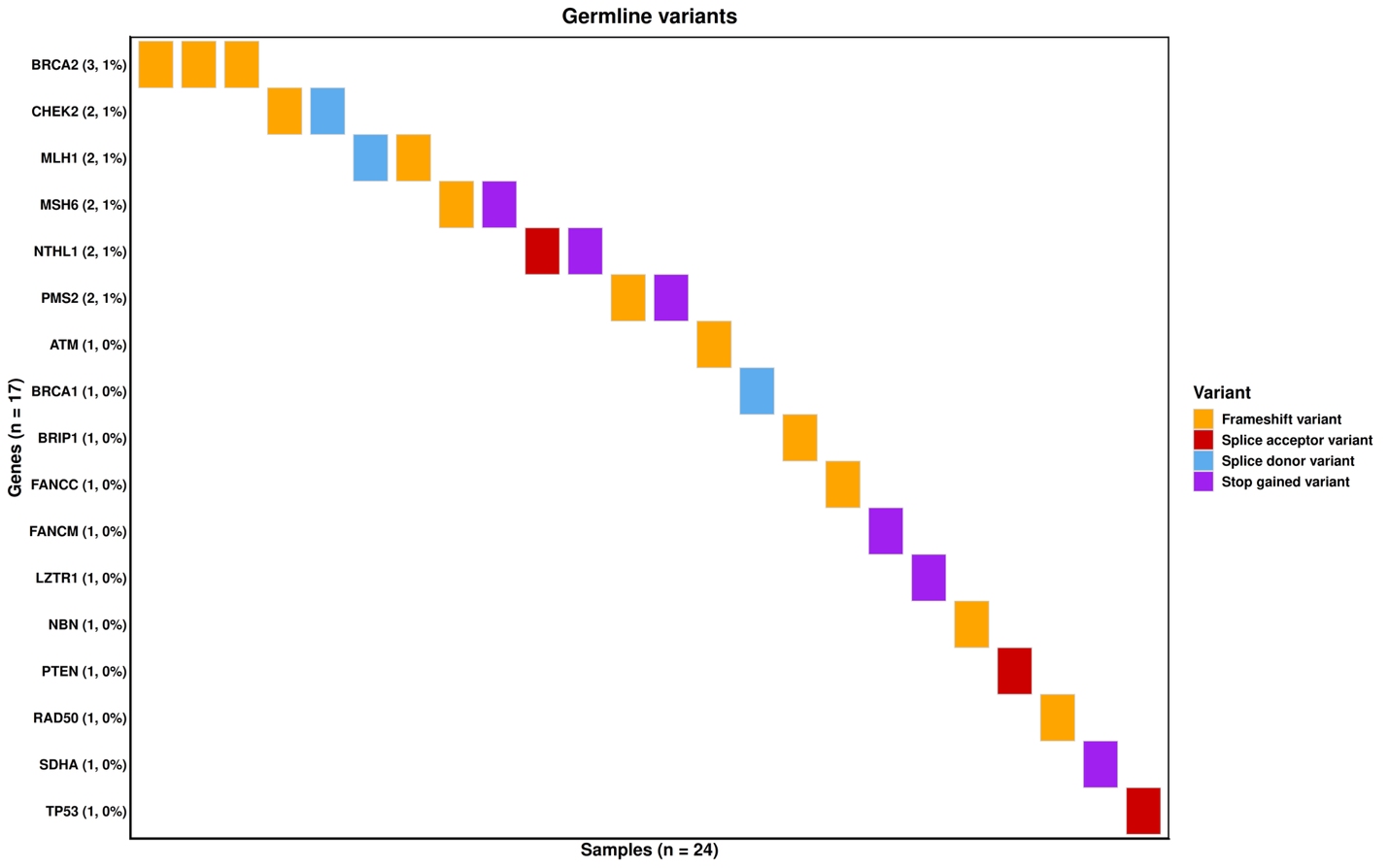

#### Supplementary Figure 5. Pathogenic or likely pathogenic germline variants.

An overview of germline variants, colored by the respective alteration (frameshift variants, splice acceptor variants, splice donor variants, and stop gained variants). A total of 24 patients (8%) had pathogenic germline variants in cancer susceptibility genes.

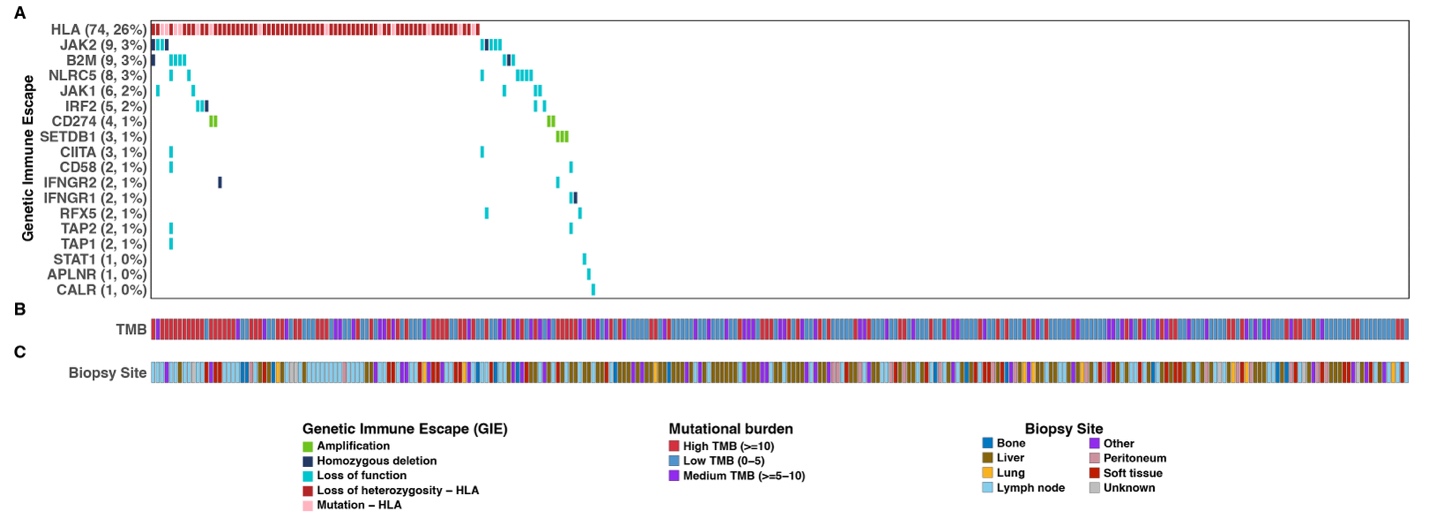

#### Supplementary Figure 6. Genetic immune escape mutations and their association with Tumor Mutational Burden (TMB) and biopsy site in CUP

(A) Genetic immune escape (GIE) mutations in genes involved in six immune-related pathways with tumor samples horizontally placed.

(B) TMB status per Mb for all samples, categorized into low(blue), medium(purple), and high (red) TMB categories.

(C) Biopsy site for all samples.

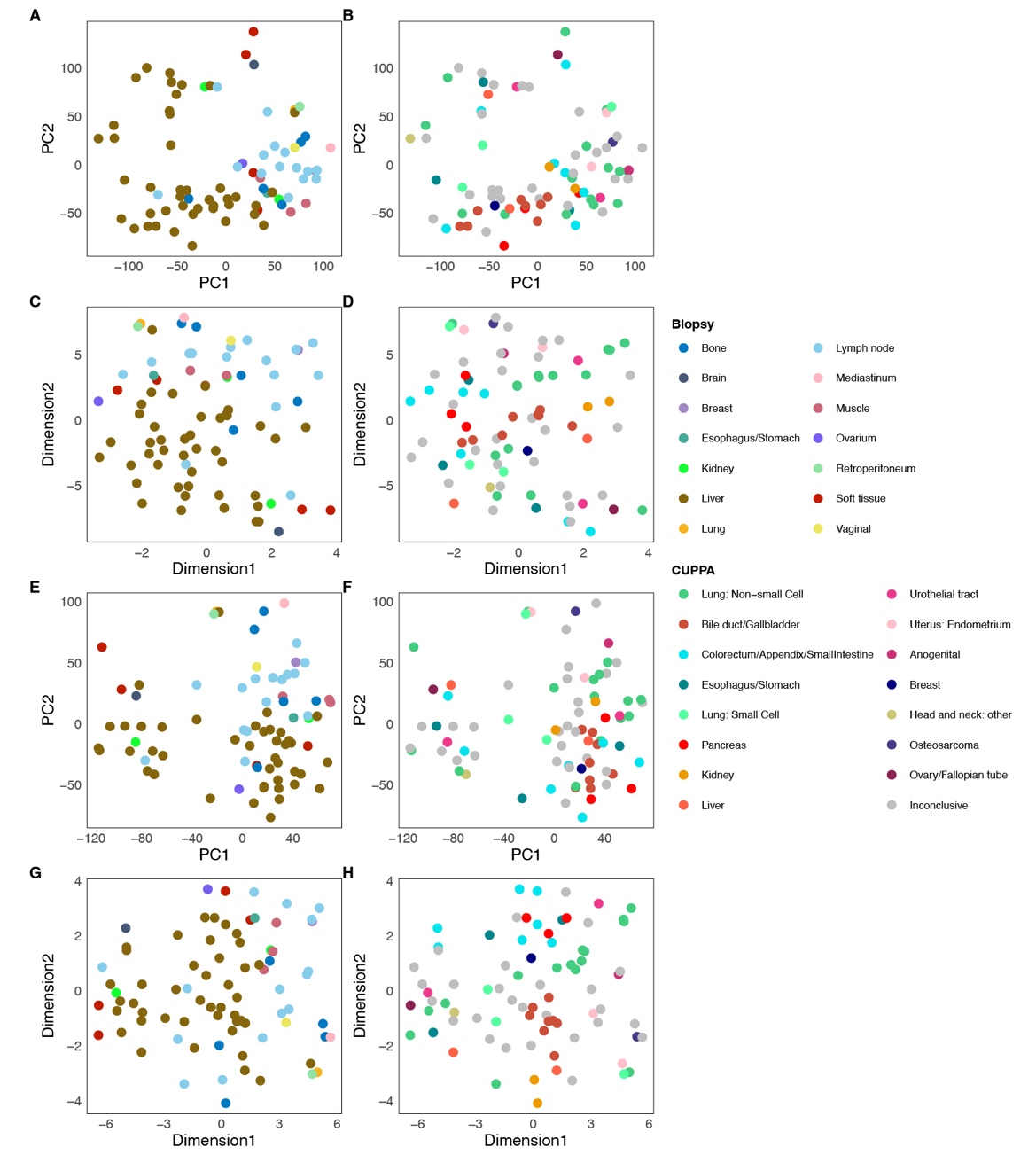

#### Supplementary Figure 7. PCA and *t*-SNE demonstrate biopsy-site effects and their reduction after gene removal

(A)-(B) PCA of RNA-seq samples using the top 30% most variable protein-coding genes, colored by biopsy site (A) and CUPPA-predicted origin (B).

(C)-(D) *t*-SNE visualization of the same data, colored by biopsy site (C) and CUPPA prediction (D).

(E)-(F) PCA after removing biopsy-enriched genes from the variable gene set, colored by biopsy site (E) and CUPPA prediction (F).

(G)-(H) *t*-SNE after biopsy-gene removal, colored by biopsy site (G) and CUPPA prediction (H).

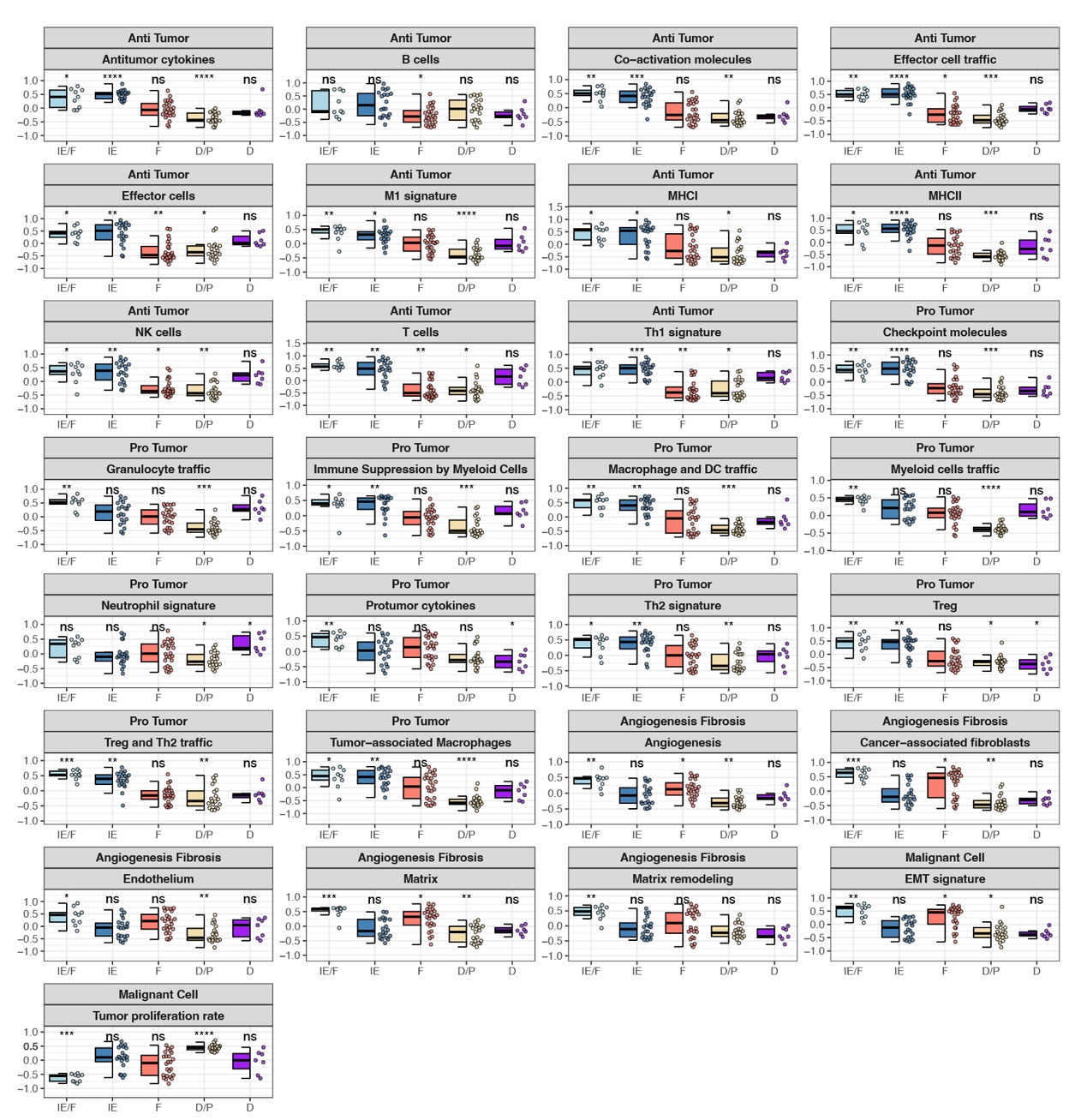

#### Supplementary Figure 8. Comparison of 29 Fges expression level across five transcriptomic subtypes

Boxplots depict the expression levels of 29 Fges across five tumor subtypes: immune-enriched/fibrotic (IE/F), immune-enriched/non-fibrotic (IE), fibrotic (F), immune-depleted/tumor proliferated (D/P), and immune-depleted (D). Fges were categorized into functional groups: anti-tumor immune cell properties, pro-tumor immune cell properties, angiogenesis/fibrosis, and malignant cell properties. Each dot represents an individual CUP. Statistical comparisons were performed using the Wilcoxon rank-sum test comparing each subtype to all others, with p-values adjusted using the Benjamini-Hochberg method. Significance is indicated as : *** p< 0.001, ** p< 0.01, *p < 0.05, ns = not significant.

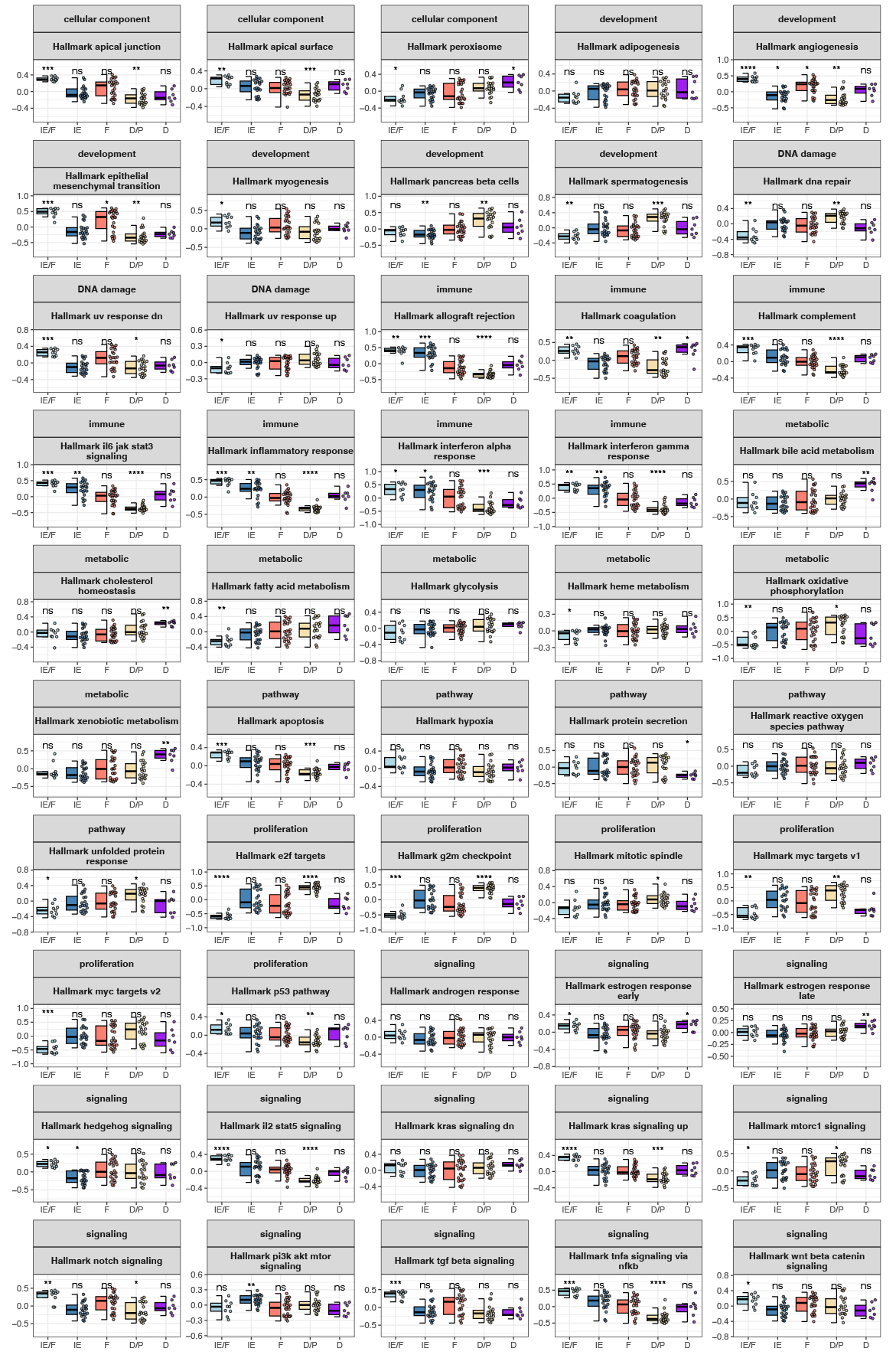

#### Supplementary Figure 9. Comparison of 50 hallmark gene set expression across 5 transcriptomic subtypes

Boxplots show expression scores for 50 hallmark gene sets, categorized into eight functional groups. Gene set expression was quantified using GSVA on RNA-seq data from CUP samples. Statistical comparisons were performed across five transcriptomic Tumor Microenvironment subtypes, using the Wilcoxon-rank sum test between each subtype and all others. P-values were adjusted using the Benjamini-Hochberg method. Significant levels denoted as follows: *** p< 0.001, ** p< 0.01, *p < 0.05, ns = not significant.

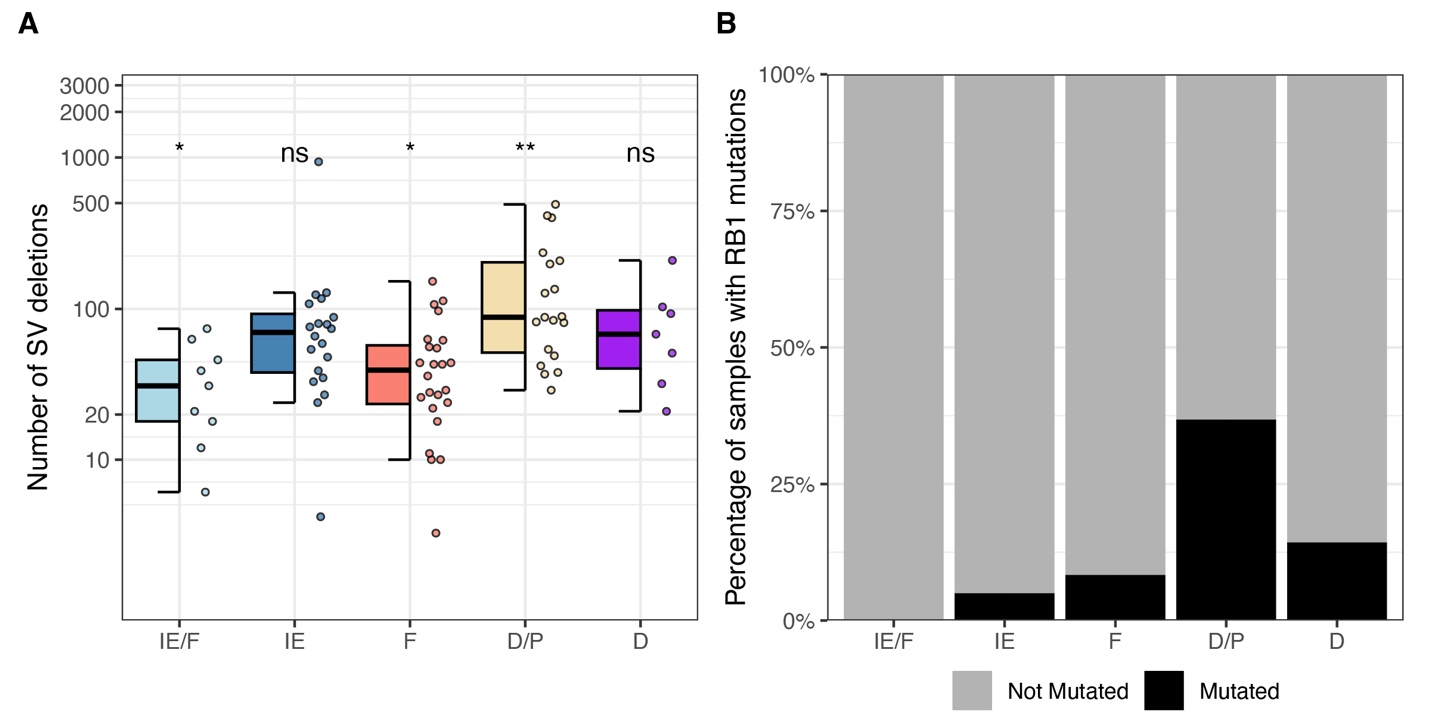

#### Supplementary Figure 10. Structural Variant distribution and *RB1* mutation status among 5 transcriptomic subtypes

(A) Number of structural deletion variants in each transcriptomic subtype. P-values were adjusted using the Benjamini-Hochberg method. Significant levels denoted as follows: *** p< 0.001, ** p< 0.01, *p < 0.05, ns = not significant.

(B) Percentage of patients with and without mutated *RB1* in each transcriptomic subtype. No significant enrichment was observed after multiple-testing correction.

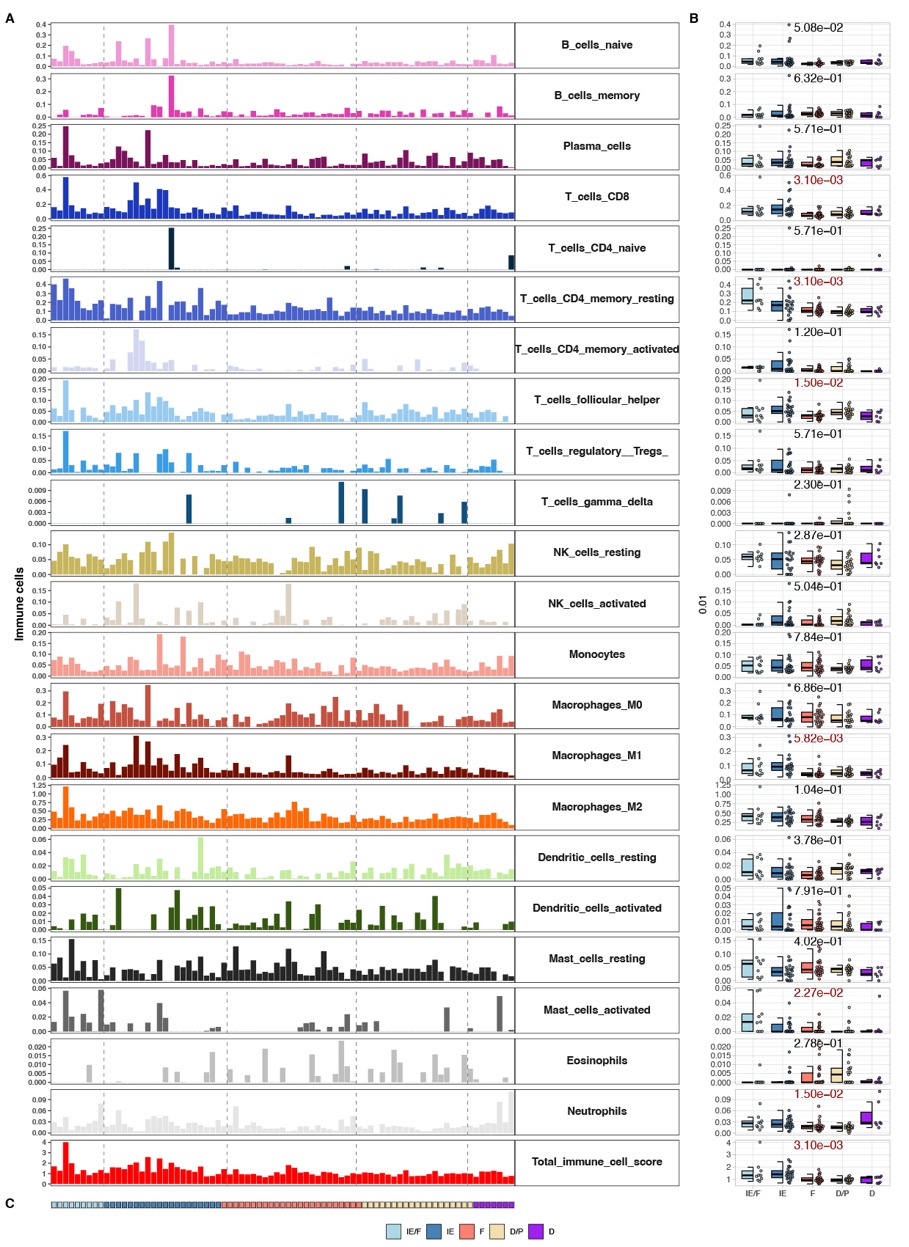

#### Supplementary Figure 11. Immune cell composition inferred by CIBERSORTx supports tumor subtype-specific tumor microenvironment (TME) characteristics in CUP

(A) Bar plots showing estimated relative abundances of 22 immune cell subtypes and a total immune cell score across CUP tumors, inferred from 79 bulk RNA-seq datasets using CIBERSORTx. Each bar represents a single CUP sample, and samples are ordered by TME subtype.

(B) Box and violin plots comparing the abundance of each immune cell type across the five TME subtypes. Statistical comparisons were performed using the Wilcoxon rank-sum test, with p-values corrected for multiple testing using the Benjamini-Hochberg method. Adjusted p-values were displayed in each panel. Significant adjusted p-values (<0.05) were marked in red.

(C) Color bar indicating the TME subtype classification of each CUP sample, aligned with panels (A) and (B)

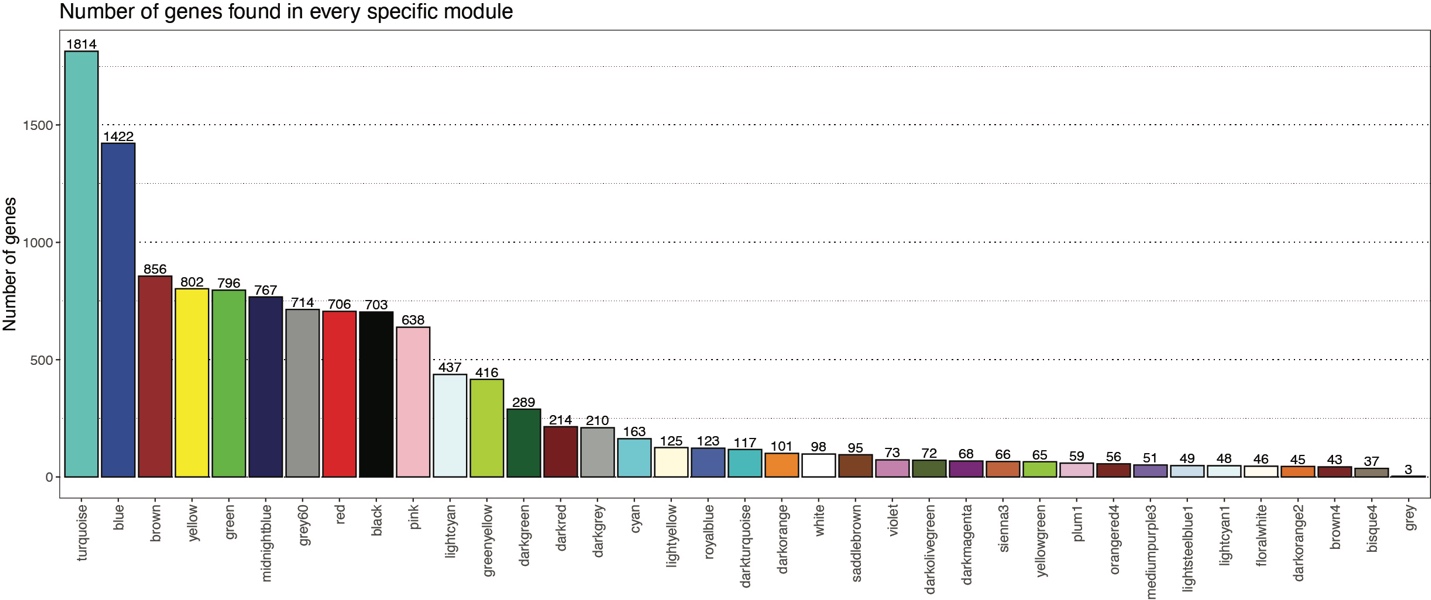

#### Supplementary Figure 12. Weighted gene expression network analysis (WGCNA) identifies 37 co-expression modules from the CUP cohort

The plot shows the number of genes per WGCNA module. The grey module contains genes that were not assigned to any co-expression module.

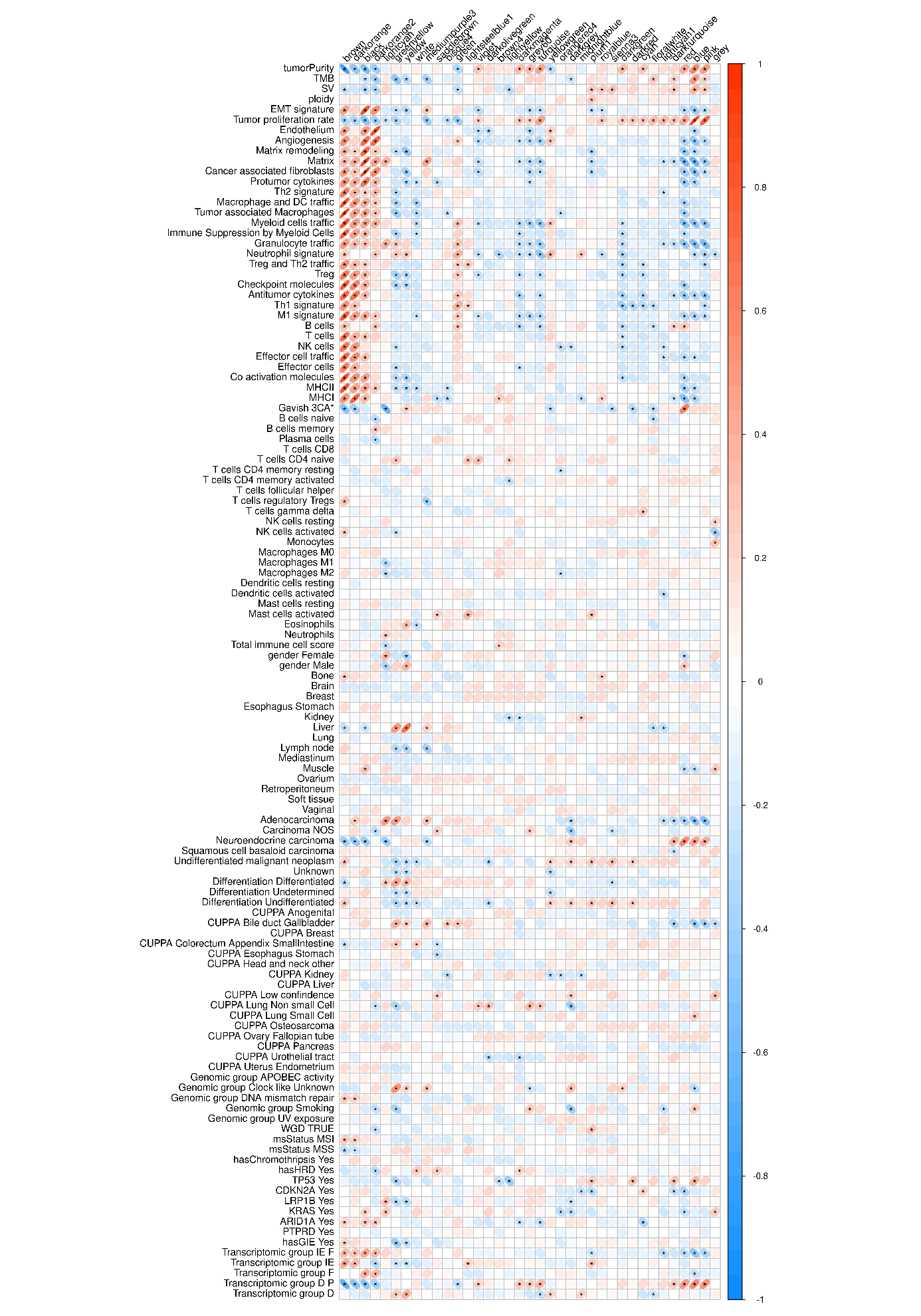

#### Supplementary Figure 13. Module-trait correlation plot across Gene expression network modules and molecular, histopathological and clinical characteristics.

Correlation plot showing all WGCNA module trait Spearman’s correlation coefficients. Ellipses represent the strength and direction of the Spearman’s correlation coefficients between module eigengenes and individual traits, with red and blue indicating positive and negative correlations, respectively. Asterisks represent statistically significant correlations (p<0.05) calculated using an asymptotic Student’s t approximation. * Gavish_3Ca_Metaprogram_Epithelial_Neuroendocrine

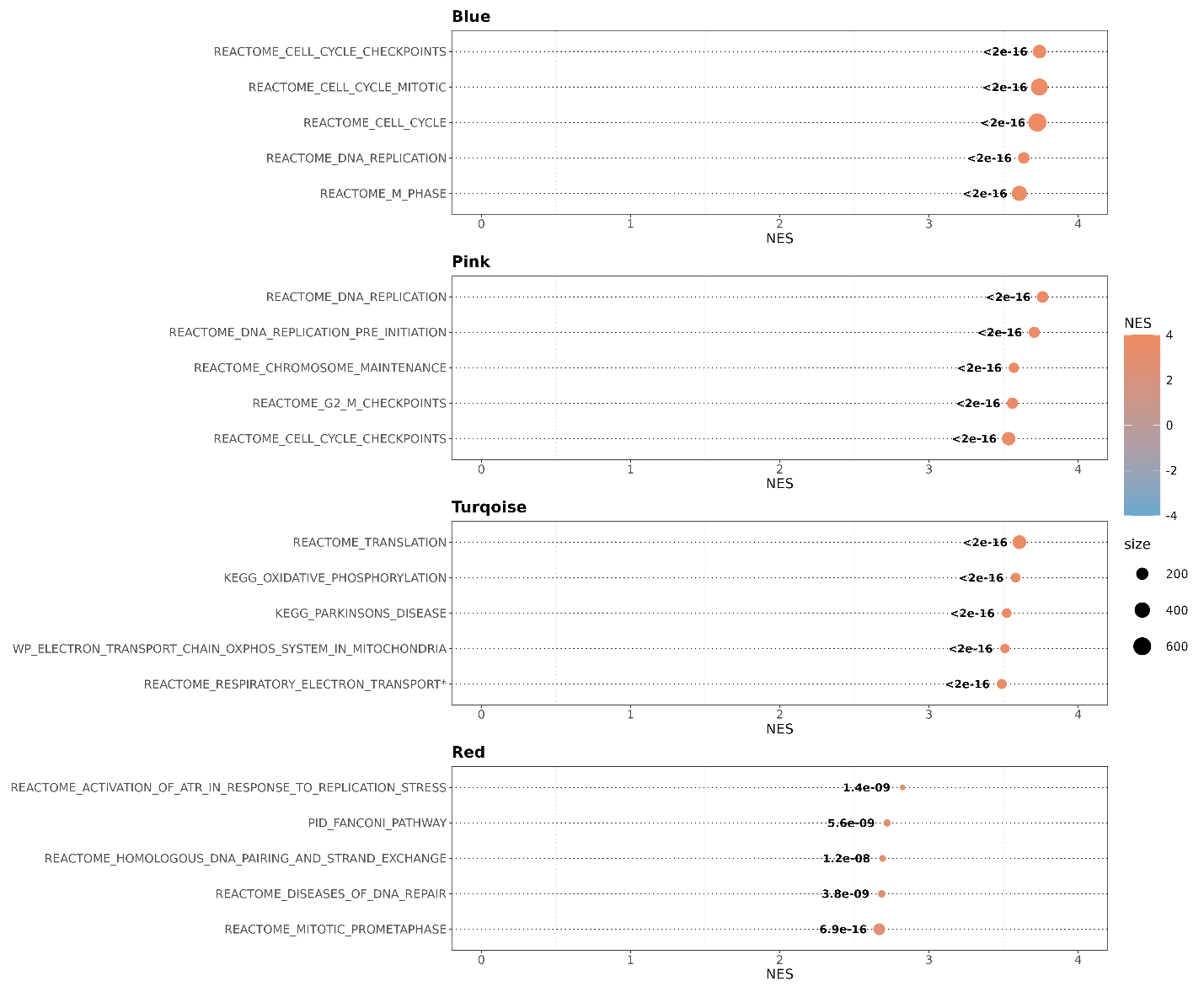

#### Supplementary Figure 14. Gene set enrichment analysis (GSEA) of selected gene expression network modules.

GSEA of gene expression network module genes using module membership, defined as the Spearman correlation between individual gene expression and the module eigengene, as ranking against the MSigDB C2: curated gene sets – CP: Canonical Pathway database. Shown p-values were calculated by fgsea and adjusted with the Benjamini-Hochberg method.

^*^Reactome_respiratory_electron_transport_atp_synthesis_by_chemiosmotic_coupling_and_heat_production_by_uncoupling_proteins

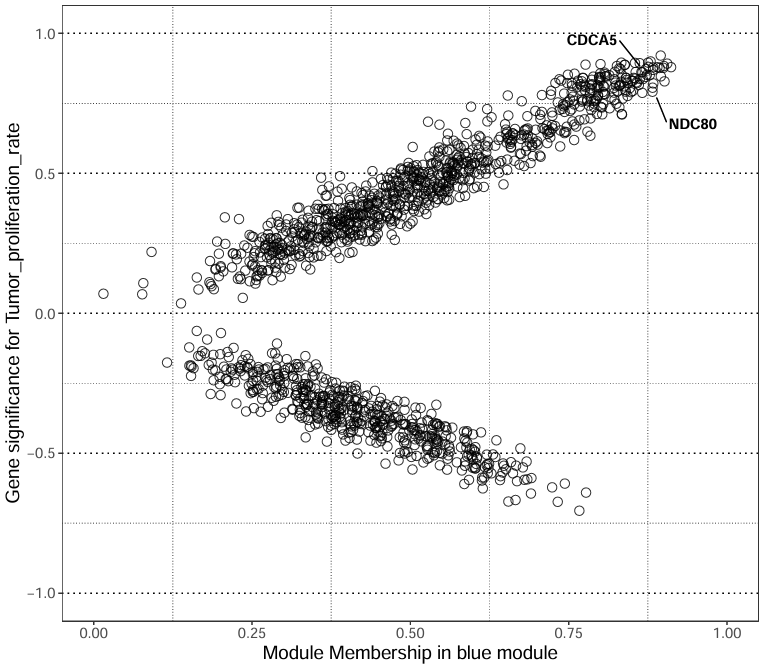

#### Supplementary Figure 15. Module membership (MM) versus gene significance (GS) for tumor proliferation rate in the blue module.

Each point represents a gene. MM is defined as the Spearman’s correlation coefficient between individual gene expression and the blue module eigengene. GS is defined as the Spearman’s correlation between individual gene expression and the tumor proliferation rate trait. CDCA5 and NDC80 are highlighted based on prior evidence from the literature.

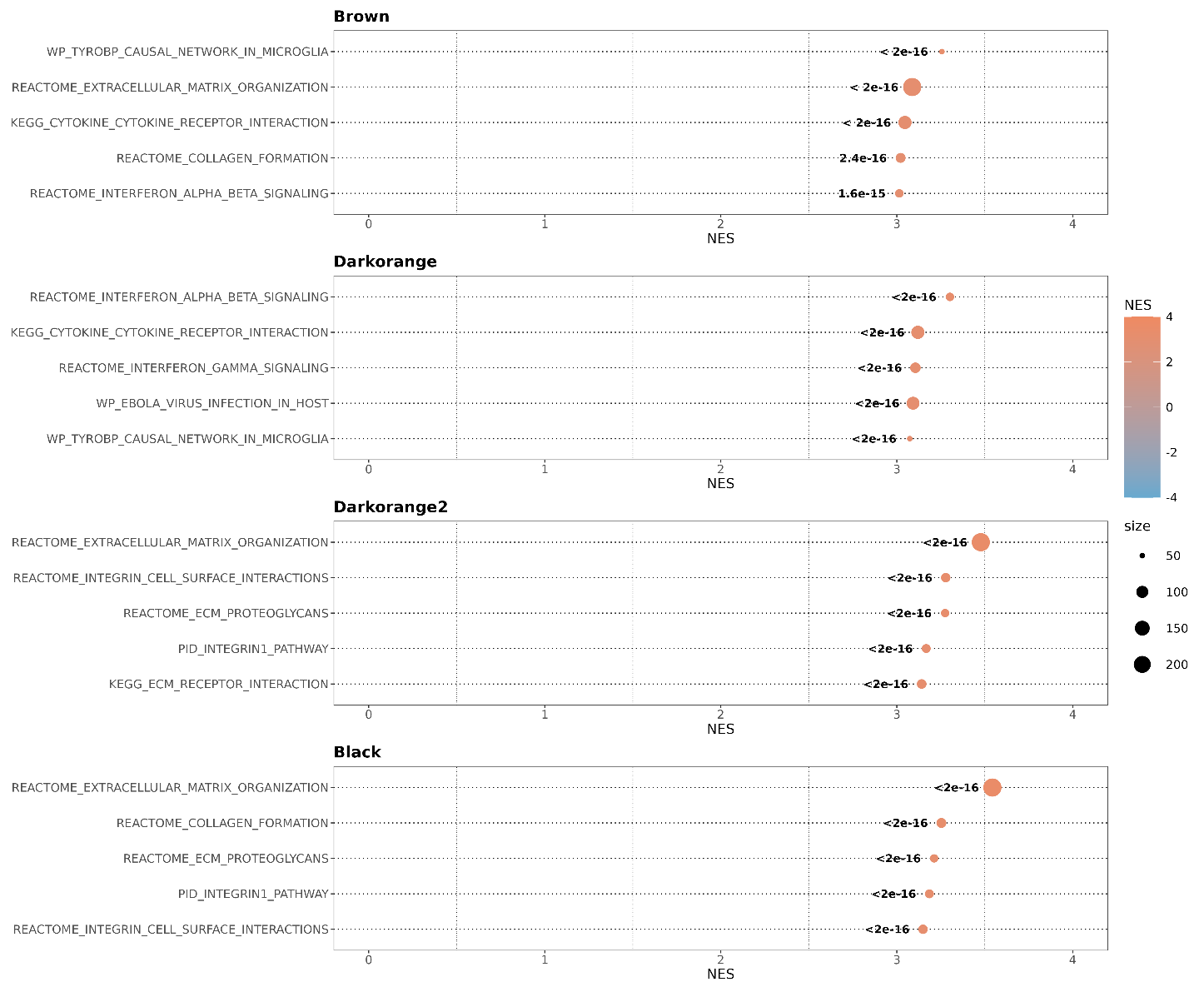

#### Supplementary Figure 16. GSEA of selected gene expression network modules.

GSEA of gene expression network module genes using module membership, defined as the Spearman’s correlation coefficient between individual gene expression and the module eigengene, as ranking against the MSigDB C2: curated gene sets – CP: Canonical Pathway database. Shown p-values were calculated by fgsea and adjusted with the Benjamini-Hochberg method.

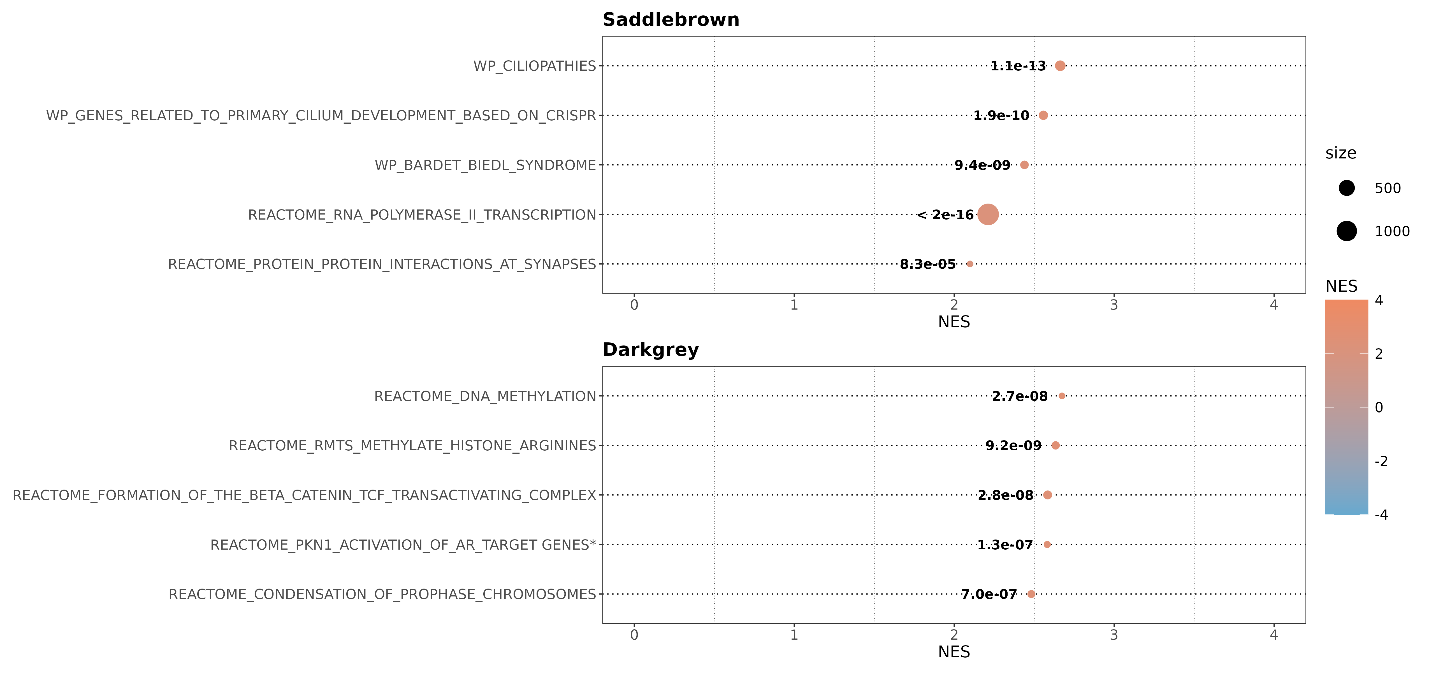

#### Supplementary Figure 17. GSEA of selected gene expression network modules.

GSEA of gene expression network module genes using module membership, defined as the Spearman’s correlation coefficient between individual gene expression and the module eigengene, as ranking against the MSigDB C2: curated gene sets – CP: Canonical Pathway database. Shown p-values were calculated by fgsea and adjusted with the Benjamini-Hochberg method. ^*^Reactome_activated_pkn1_stimulates_transcription_of_ar_androgen_receptor_regulated_genes_klk2_and_klk3

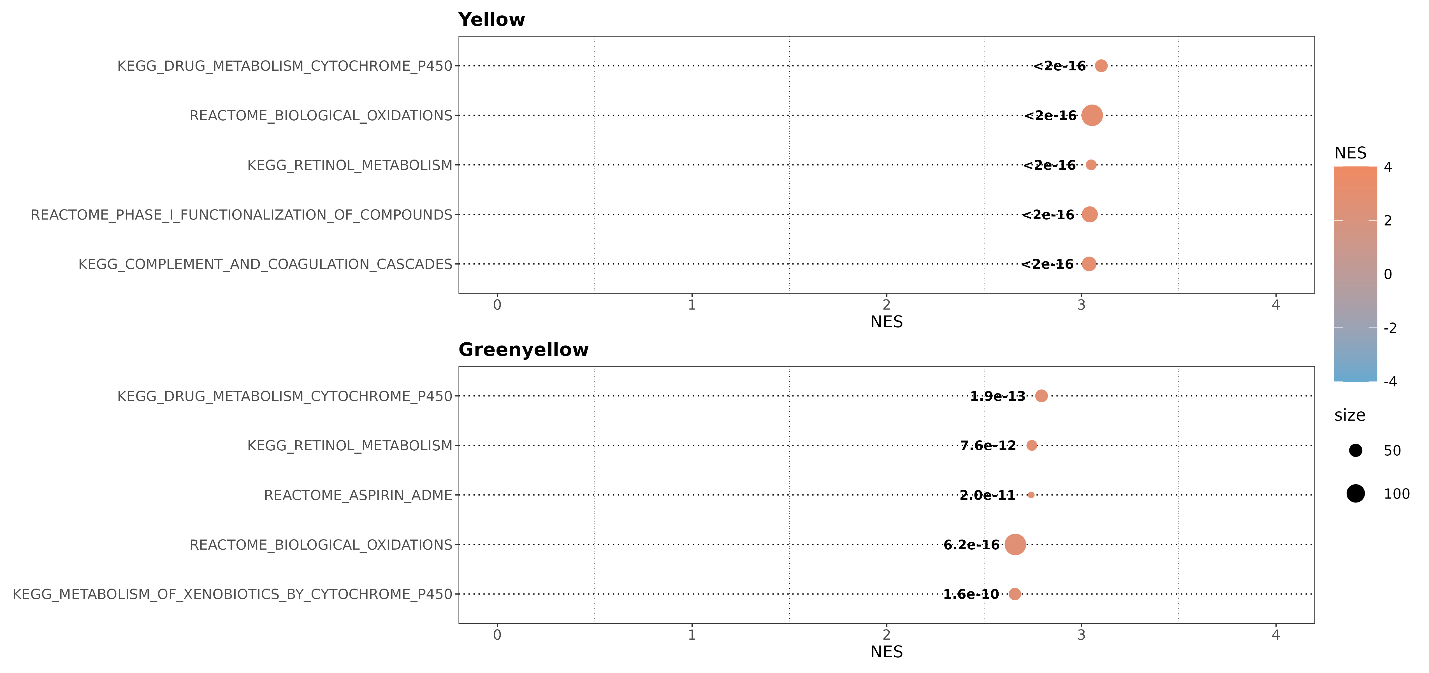

#### Supplementary Figure 18. GSEA of selected gene expression network modules.

GSEA of gene expression network module genes using module membership, defined as the Spearman correlation between individual gene expression and the module eigengene, as ranking against the MSigDB C2: curated gene sets – CP: Canonical Pathway database. Shown p-values were calculated by fgsea and adjusted with the Benjamini-Hochberg method.
